# Large-scale genetic characterization of Parkinson’s disease in the African and African admixed populations

**DOI:** 10.1101/2025.01.14.25320205

**Authors:** Fulya Akçimen, Kimberly Paquette, Peter Wild Crea, Paula Saffie-Awad, Charles Achoru, Funmilola Taiwo, Simon Ozomma, Gerald Onwuegbuzie, Marzieh Khani, Spencer Grant, Lukman Owolabi, Chiamaka Okereke, Olajumoke Oshinaike, Emmanuel Iwuozo, Paul Suhwan Lee, Shyngle Oyakhire, Nosakhare Osemwegie, Kensuke Daida, Sani Abubakar, Adedunni Olusanya, Mariam Isayan, Rami Traurig, Adebimpe Ogunmodede, Sarah Samuel, Mary B. Makarious, Fadimatu Sa’ad, Rashidat Olanigan, Kristin Levine, Ewere Marie Ogbimi, Dan Vitale, Francis Odiase, Mathew J. Koretsky, Francis Ojini, Olanike Odeniyi, Zih-Hua Fang, Nkechi Obianozie, Deborah A. Hall, Ernest Nwazor, Tao Xie, Francisca Nwaokorie, Mahesh Padmanaban, Paul Nwani, Ejaz A. Shamim, Alero Nnama, David Standaert, Morenikeji Komolafe, Marissa Dean, Godwin Osaigbovo, Elizabeth Disbrow, Ismaila Ishola, Ashley Rawls, Frank Imarhiagbe, Shivika Chandra, Cyril Erameh, Vanessa Hinson, Naomi Louie, Ahmed Idowu, J Solle, Scott A. Norris, Abdullahi Ibrahim, Camilla Kilbane, Gauthaman Sukumar, Lisa M. Shulman, Daniel Ezuduemoih, Julia Staisch, Sarah Breaux, Clifton Dalgard, Erin R. Foster, Abiodun Bello, Andrew Ameri, Raquel Real, Erica Ikwenu, Huw R Morris, Roosevelt Anyanwu, Erin Furr Stimming, Kimberley Billingsley, Wemimo Alaofin, Pilar Alvarez Jerez, Osigwe Agabi, Dena G. Hernandez, Rufus Akinyemi, Sampath Arepalli, Laksh Malik, Raymond Owolabi, Yakub Nyandaiti, Hampton L. Leonard, Kolawole Wahab, Kathryn Step, Oladunni Abiodun, Carlos F. Hernandez, Fatima Abdulai, Hirotaka Iwaki, Soraya Bardien, Christine Klein, John Hardy, Henry Houlden, Kamalini Ghosh Galvelis, Mike A. Nalls, Nabila Dahodwala, Whitley Aamodt, Emily Hill, Alberto Espay, Stewart Factor, Chantale Branson, Cornelis Blauwendraat, Andrew B. Singleton, Oluwadamilola Ojo, Lana M. Chahine, the Black and African American Connections to Parkinson’s Disease Study (BLAAC PD), the Nigeria Parkinson’s Disease Research Network (NPDRN), the Racial Disparities in Parkinson Disease (RaD-PD) and the Global Parkinson’s Genetics Program (GP2), Njideka Okubadejo, Sara Bandres-Ciga

## Abstract

Elucidating the genetic contributions to Parkinson’s disease (PD) etiology across diverse ancestries is a critical priority for the development of targeted therapies in a global context. We conducted the largest sequencing characterization of potentially disease-causing, protein-altering and splicing mutations in 710 cases and 11,827 controls from genetically predicted African or African admixed ancestries. We explored copy number variants (CNVs) and runs of homozygosity (ROHs) in prioritized early onset and familial cases. Our study identified rare *GBA1* coding variants to be the most frequent mutations among PD patients, with a frequency of 4% in our case cohort. Out of the 18 *GBA1* variants identified, ten were previously classified as pathogenic or likely pathogenic, four were novel, and four were reported as of uncertain clinical significance. The most common known disease-associated *GBA1* variants in the Ashkenazi Jewish and European populations, p.Asn409Ser, p.Leu483Pro, p.Thr408Met, and p.Glu365Lys, were not identified among the screened PD cases of African and African admixed ancestry. Similarly, the European and Asian *LRRK2* disease-causing mutational spectrum, including *LRRK2* p.Gly2019Ser and p.Gly2385Arg genetic risk factors, did not appear to play a major role in PD etiology among West African-ancestry populations. However, we found three heterozygous novel missense *LRRK2* variants of uncertain significance overrepresented in cases, two of which — p.Glu268Ala and p.Arg1538Cys — had a higher prevalence in the African ancestry population reference datasets. Structural variant analyses revealed the presence of *PRKN* CNVs with a frequency of 0.7% in African and African admixed cases, with 66% of CNVs detected being compound heterozygous or homozygous in early-onset cases, providing further insights into the genetic underpinnings in early-onset juvenile PD in these populations. Novel genetic variation overrepresented in cases versus controls among screened genes warrants further replication and functional prioritization to unravel their pathogenic potential. Here, we created the most comprehensive genetic catalog of both known and novel coding and splicing variants potentially linked to PD etiology in an underserved population. Our study has the potential to guide the development of targeted therapies in the emerging era of precision medicine. By expanding genetics research to involve underrepresented populations, we hope that future PD treatments are not only effective but also inclusive, addressing the needs of diverse ancestral groups.

## Introduction

Parkinson’s disease (PD) is a multifaceted neurodegenerative disorder influenced by genetics, environment, and other factors. The influence of genetics has been primarily studied in European populations, limiting our understanding of the genetic landscape of PD in the African and African admixed populations ^1^. Bridging this gap is essential for advancing equitable precision medicine interventions and developing universally effective strategies for the prevention and treatment of PD.

Genetic risk factors and their impact on PD in individuals of African and African admixed genetic ancestry remain largely unknown. Around 77% of African genetic studies in PD have been conducted in North Africa (mainly Tunisia) and South Africa ^2^. In contrast, there have been limited PD genetic studies in populations originating from Central, Eastern, and the French speaking West coast of Africa. Similarly, the genetics of African admixed individuals have been poorly studied in the context of PD genetics, with few published studies ^3–5^. As a result, our understanding of the role of genetics in disease etiology among African admixed individuals lags behind that of European ancestry individuals. Further research with more diverse representation and standardization is crucial for drawing accurate conclusions on a global scale.

The first Genome-Wide Association Study (GWAS) exploring PD genetic risk in African and African admixed populations nominated a novel non-coding PD-associated variant (rs3115534) in *GBA1* ^6^. This variant is present in 33% and 22% of the African and African admixed PD cohorts respectively ^6^ while it is almost completely absent in predominantly European ancestry cohorts. The rs3115534 intronic variant acts by interfering with the splicing of functional *GBA1* transcripts, resulting in reduced glucocerebrosidase activity. This represents a novel mechanism of *GBA1*-derived PD risk and an attractive candidate for precision-based therapeutics in a remarkably underserved population ^7^, reinforcing the idea that distinct genetic architectures contribute to disease susceptibility and in turn could inform optimized treatment options in the future. Other *GBA1* coding variants, like p.Asn409Ser and p.Leu483Pro, are associated with PD risk and commonly observed in European and Ashkenazi Jewish ancestry populations. These same variants are notably rare among individuals of African and African admixed ancestries ^8,9^.

To gain a deeper understanding of the genetic architecture of PD in underserved African populations, we leveraged data from the Black African and African American Connections to Parkinson’s Disease Study (BLAAC PD) ^10^ as a part of the Global Parkinson’s Genetics Program (GP2) ^11^, the Nigerian Parkinson’s Disease Research (NPDR) Network ^12^, the PDGENEration (PD GENE) initiative ^13^, All of Us ^14^, and the UK Biobank (UKB) ^15^. Utilizing data from 710 cases and 11,827 controls, we conducted the largest sequencing characterization of potentially disease-causing, protein-altering, and splicing mutations across 51 genes in African and African admixed ancestry populations. We focused on known PD-associated genes linked to autosomal dominant inheritance (*SNCA, LRRK2* and *VPS35*), autosomal recessive inheritance (*PRKN, PINK1, DJ-1,* and *VPS13C*), and increased risk (*LRRK2, GBA1*), as well as other PD-associated genes such as *RAB32*, *PSMF1*, and *ITSN1* from literature. Furthermore, we assessed putative pleiotropic effects by examining genes that have been shown to manifest as parkinsonism ^16^ (i.e. *ATP13A2*, *FBXO7, PLA2G6, SYNJ1, GCH1*, *TRPM7, GCH1, KCDT7*, *XPR1,* and *TOR1A*) or other neurodegenerative conditions (i.e. *APP, MAPT*, and *DCTN1*) (**Supplementary table 1**). We leveraged available short-read sequencing datasets (whole genome, whole exome, and targeted exome sequencing) to comprehensively unravel the genetic architecture of rare variants contributing to PD in these populations. We further investigated potential recessive variants by exploring runs of homozygosity (ROHs) enriched in the patient population. We assessed potential copy number variation (CNVs) by investigating B allele frequency (BAF) and Log2 Ratio (L2R) from genotyping data along with Multiplex ligand-dependent probe amplification (MLPA), focusing on cases with positive family history and early age at onset.

## Methods

### Participants and study design

We obtained genetic data from All of Us, GP2-BLAAC PD, the NPDR Network, PD GENE, and UKB datasets **(Table 1)**. In total, the data comprised 710 cases and 11,827 controls of African or African admixed ancestry or Black or African American self-reported race, See **Figure 1** for demographic characteristics of cohorts under study. See **Figure 2** for a summary of our workflow, explained in further detail below.

BLAAC PD is a multicenter study in North America recruiting individuals who self-identify as Black and/or African American. In the GP2-BLAAC PD dataset, cases were defined as individuals aged 18 years or older in the United States with a clinician-confirmed PD diagnosis (n=186), whereas controls were neurologically healthy individuals with no personal or family history of any neurodegenerative disease (n=236). Institutional review board (IRB) approval was implemented at each study site, with a written informed consent provided by each participant ^17^. Assessments to determine cognitive status and clinical features were conducted and reported by the respective clinicians. Cognitive status was determined by the cognition component of the Clinical Impression of Severity Index for PD (CISI-PD) with scores ranging from 0 to 6: 0 (normal), 1 (slowness and/or minimal cognitive problems), 2 (mild cognitive problems; no limitations), 3 (mild to moderate cognitive problems; does not need help for basic activities of daily living), 4 (moderate cognitive problems; help is required for some basic activities of daily living), 5 (severe cognitive problems; help is required for most or all basic activities of daily living), and 6 (severely disabled; helpless; complete assistance needed) ^18^. The occurrence of clinical symptoms such as bradykinesia, rest tremor, rigidity, and use/response to levodopa was assessed through the International Parkinson and Movement Disorder Society (MDS) Clinical Diagnostic Criteria for idiopathic PD (MDS-PD criteria) ^19^.

For the NPDR Network cohort (n cases =108; n controls=60), PD diagnosis was based on the United Kingdom PD Society Brain Bank criteria (excluding the requirement for not more than one affected relative) ^12,20^. Study assessments include the Movement Disorders Society Unified Parkinson’s Disease Rating Scale (MDS-UPDRS), a one question, self-reported olfactory assessment, and a neurological examination, performed by a study neurologist. Controls were excluded if any clinical signs of neurological disease were present ^12^. All participants provided written informed consent.

In addition, we mined existing data from PD GENE, a multi-center, observational study (NCT04057794, NCT04994015) offering CLIA-certified genetic testing and genetic counseling to individuals with PD in North America ^13^. We included targeted exome sequencing data from 138 PD cases with genetically predicted African or African admixed ancestry and 145 self-reported Black or African American PD cases from the GP2 release 8. The study was approved by IRBs, as well as the Scientific Review and Executive Committees of the Parkinson Study Group. All participants signed informed consent forms.

In the All of Us dataset, we created a case-control cohort comprising 93 PD cases and 10,360 controls of African or African admixed ancestry. Controls were individuals aged 60 years or older with no family history of neurological disease and no present neurological condition in their electronic health records. All participants provided written informed consent. The All of Us IRB reviews the protocol, informed consent, and other materials for All of Us participants.

PD cases of genetically predicted African ancestry in the UKB (n=40) were defined by the UKB field ID 42032, using diagnoses according to the UKB’s algorithmically defined outcomes v2.0 (https://biobank.ndph.ox.ac.uk/ukb/refer.cgi?id=460). The control cohort (n=1,171) includes individuals aged 60 years or older without any neurological condition or family history of any neurological disorders.

Sequencing, genotyping procedures, and quality assessment of 710 cases and 11,827 controls are detailed in Supplementary Methods.

### Assessment of relatedness

Pairwise relatedness inference was estimated using the KING v.2.3.1 (Kinship-based INference for GWAS) software ^21^. In brief, samples with a Kinship coefficient of at least 0.0884 were further explored for relatedness. Related samples are inferred based on the range of estimated kinship coefficients: >0.354, 0.354-0.177, 0.177-0.0884, and 0.0884-0.0442 that correspond to duplicate/monozygotic twin, 1st-degree, 2nd-degree, and 3rd-degree relationships, respectively.

### Runs of homozygosity

To investigate genome-wide homozygosity, runs of homozygosity (ROHs) were identified using PLINK v.1.9 ^22^, as described below. A sliding window of 50 SNPs (--homozyg-window-snp) and 1,500kb in length (--homozyg-kb) was used in a stepwise approach as previously described ^23^. A minimum of 100 SNPs (--homozyg-snp) was allowed with a threshold of one heterozygous SNP (--homozyg-window-het) and five missing SNPs (--homozyg-window-missing) in a ROH region. To enable the detection of shorter ROHs, a ROH region was called if each SNP covered a minimum of 5% of the homozygous sliding window (--homozyg-window-threshold). The ROH cut-off was set at >1.5Mb (--homozyg-kb), with a maximum allowable distance of 1Mb between consecutive SNPs (--homozyg-gap) and a minimum density requirement of one SNP per 50kb of the genome (--homozyg-density). Furthermore, we investigated ROH intersecting with known PD genes and GWAS loci as well as parkinsonism gene regions. Here, we used an approximate 1MB window both upstream and downstream from the known genes and GWAS hits ^6,24–27^.

### Copy number variant detection

CNV detection was performed leveraging GP2-BLAAC PD genotyping data by assessing B allele frequency (BAF) and Log2 Ratio (L2R) estimates for 1,167 PD cases (African =872, African admixed =295). Additional details are highlighted in Supplementary Methods. Briefly, L2R ratio gives an indirect measure of the copy number of each SNP by plotting the ratio of observed to expected hybridization intensity. An R value above 1 is indicative of an increase in copy number (duplication or triplication), and an R value below 1 suggests a decrease (deletion). BAF plots the proportion of times an allele is called A or B at each genotype: thus the expected ratios are 1.0 (B/B), 0.5 (A/B) and 0.0 (A/A). Significant deviations from these figures in contiguous SNPs are indicative of a CNV. While this metric exhibits a high level of variance for individual SNPs, it does provide a measure of CNV when L2R values for numerous adjacent SNPs are visualized as described elsewhere ^28^. BAF and L2R were extracted for *SNCA* and *PRKN* with ± 250 kb proximity of the target genes as these genes are particularly enriched for genetic rearrangements in PD.

### Genetic characterization of potentially rare disease-causing variants

We performed functional annotation of identified variants using ANNOVAR ^29^, incorporating relevant datasets such as refGene, dbnsfp47a, which incorporates the AlphaMissense and PrimateAI prediction tools, and gnomAD v4.1. To distinguish *GBA1* variation from *GBAP1* pseudogene mutations, we implemented the Gauchian targeted caller ^30^ on short-read whole genome sequencing (WGS) data to detect known *GBAP1* variation within the exons 9–11 homology region. The clinical significance of candidate variants was cross-checked on the ClinVar database (https://www.ncbi.nlm.nih.gov/clinvar/). Genes implicated in parkinsonism and other related neurodegenerative diseases were annotated using the OMIM genemap2 file (downloaded from omim.org in September 2024). To identify rare, potentially disease-causing mutations, we focused on missense, loss-of-function variants including stopgain, stop loss, frameshift, start loss, splice site changes, and short insertions and deletions, with a minor allele frequency (MAF) below 0.01 and a minimum Combined Annotation Dependent Depletion (CADD) score of 12.37, which is predicted to be among the 2% most pathogenic variants of the genome, as suggested by Kircher et al. ^31^. We assigned a confidence level for PD genes using the criteria described by Blauwendraat, et al. ^32^.

The predicted protein structures encoded by eight genes associated with PD (*GBA1*, *LRRK2*, *DJ-1, PINK1*, *PRKN*, *SNCA*, *VPS13C*, *VPS35*) were obtained from the EMBL AlphaFold Protein Structure Database to ensure that all of the residues are present in each protein structure ^33,34^. PyMOL v. 2.6.0 was used to represent the protein structures and their associated mutation sites from identified genetic variants (The PyMOL Molecular Graphics System, Version 2.6.0 Schrödinger, LLC).

## Results

### Baseline characteristics

An overview of our study is illustrated in **Figure 1**. Demographic and clinical characteristics of five datasets including 710 cases and 11,827 neurological healthy controls are summarized in **Table 1**. All participants were of African and African admixed ancestry based on genetic prediction, except for the PD GENE dataset, where 138 participants genotyped on the NeuroBooster array were confirmed as African or African admixed, and 145 participants were included based on self-reported Black or African American race **(Supplementary** Figure 1).

### Genetic findings

The current study identified 443 rare variants, including 105 in eight PD genes and 338 in genes associated with parkinsonism or other neurodegenerative diseases (**Supplementary Table 1**). A total of 19 variants were previously reported as pathogenic or likely pathogenic in ClinVar, while 182 variants were novel. A total of 242 variants were reported as benign, likely benign or of uncertain clinical significance. The ClinVar dataset is largely based on mutation reports from European populations and so is likely to have incomplete information on pathogenicity in African individuals. Variants were prioritized based on clinical relevance, zygosity, and the presence of an early disease onset in cases involving recessive PD genes. This process identified 84 variants across 14% of the cases (n=97) that had no prior classification as benign **(Supplementary Table 2**).

The average age at onset (AAO) for cases in our study ranged from 56.34 ± 11.90 years (NPDRN) to 66.79 ± 9.55 years (UKB). Among carriers of prioritized variants, the AAO was notably younger, at 52.41 ± 13.65 years. Furthermore, carriers with a family history of PD had an earlier AAO of 44.43 ± 13.77 years, compared to 53.64 ± 13.73 years for those without a family history. The distribution of variants in PD genes across the five datasets, excluding those previously reported as benign or likely benign, is presented in **Figure 3**.

Notably, ten variants in *GBA1*, one in *PINK1*, two in *PRKN*, one in *FBXO7,* and one in *PLA2G6* were previously described as pathogenic or likely pathogenic **(Supplementary Table 2, Figure 4).** A total of 41 novel variants were identified in PD genes, including four in *GBA1*, seven in *PINK1*, two in *DNAJC6*, four in *DJ-1*, one in *SNCA*, five in *PRKN,* three in *LRRK2*, twelve in *VPS13C*, two in *VPS35*, and one in *SYNJ1* **(Supplementary Table 2, Figure 4)**. We obtained the pathogenicity scores of the candidate variants from CADD, AlphaMissense, and PrimateAI. A score >12.36 for CADD, >0.564 for AlphaMissense, and >0.803 for PrimateAI are recommended as pathogenic cutoffs. None of the previously reported pathogenic variants in known genes fulfilled the pathogenicity criteria for AlphaMissense and PrimateAI. Therefore, although we included scores from these tools, we did not apply any filtering based on the recommended thresholds. All identified single nucleotide variants in recessive PD genes were present in single heterozygous states **(Supplementary Table 2, Figures 3 and 4**). A female PD case from the PD GENE study with an AAO under 30 was identified as a carrier of two heterozygous variants in *PRKN—* p.Gly139Valfs*38 and p.Pro113Thrfs*51*—*that would suggest a potential compound heterozygous state **(Table 2, Supplementary Table 3).** We were unable to confirm compound heterozygosity due to the absence of phasing data. A distribution of AAO among prioritized variant carriers in PD genes (*PRKN, DJ-1, PINK1, VPS13C, DNAJC6, GBA1, SNCA, LRRK2, TRPM7,* and *VPS35*), is shown in **Figure 5**. The identified genetic variants in the proteins encoded by the eight screened genes associated with PD in this study are displayed in **Figure 6,** in addition to the variants found among 11 PD or parkinsonism genes shown in **Supplementary Figure 2**.

#### Rare *GBA1* variants are the most commonly identified mutations in Parkinson’s disease patients of African and African admixed ancestry, exhibiting a distinct mutation landscape compared to other populations

We identified a total of 18 heterozygous *GBA1* variants among 26 cases (4%), including ten previously reported as pathogenic or likely pathogenic, four novel variants, and four variants of uncertain clinical significance **(Table 2, Supplementary Table 2, Figures 3,4, & Supplementary** Figure 2). To identify potential known variants in the pseudogene paralog *GBAP1* exons 9-11 homology region, we screened 15 short-read WGS samples. The Gauchian algorithm predicted four carrier mutations, including variants p.Asp448His, c.1263del and c.1263del+Rec*TL*, i.e. deletion combined with pseudogene variant in exon 9 and three in exon 10. The algorithm predicted no structural variants: a total of four copies per *GBA1* and *GBAP1* variant in each sample were identified. Overall, four variants, including p.Gly416Ser, p.Trp223Arg, p.Arg170Leu, and p.Thr75del in *GBA1* were targeted outside the homology region.

The previously reported pathogenic p.Arg170Leu (rs80356763) variant was identified in a family with two early-onset female PD siblings of African genetic ancestry, with ages at onset in their early 30s and 40s. The p.Thr75del (rs761621516) variant was identified in two cases from PD GENE and one case from the GP2-BLAAC PD dataset with no familial history. The carriers from the PD GENE were two Black or African/African American male patients (one with confirmed African genetic ancestry), who had onset in their 50s and early 70s. The carrier from the GP2-BLAAC PD dataset was a female patient with a disease onset in her late 50s. Both of these variants appear to be specific to African and African admixed ancestries, with no records in other ancestries according to gnomAD v4.1.0 and All of Us release 7 browsers **(Supplementary Table 2).**

Four additional known pathogenic *GBA1* variants, including p.Arg535His (1:155235002:C:T, rs75822236), p.Asp448His (1:155235727:C:G, rs1064651), p.Arg502Cys (1:155235196:G:A, rs80356771), and p.Asn227Ser (1:155238215:T:C, rs364897), were identified in patients from the GP2- BLAAC PD release 8 data (**Table 2**). The p.Arg535His variant was identified in a female individual of African ancestry with disease onset in her 70s, whereas the p.Asp448His and p.Arg502Cys variants were identified in female individuals of African admixed ancestry with onset in their 50s. The p.Asn227Ser variant was identified in a male patient of African ancestry with onset in his early 50s.

Two previously reported pathogenic variants, p.Ala495Pro (1:155235217:C:G, rs368060) and p.Gly416Ser (1:155235823:C:T, rs121908311), were found in two female patients in the All of Us dataset (**Table 2**). Both patients were of African ancestry and diagnosed in their 50s. Additionally, the known missense p.Trp223Arg (1:155238228:A:G, rs61748906) variant was identified in a male patient of African genetic ancestry from the NPDR Network cohort. He presented with initial symptoms in his early 60s and reported no family history. The stopgain p.Arg202Ter (1:155238291:G:A, rs1009850780) variant was found in a female patient from the PD GENE dataset, with onset in her late 50s (**Table 2**).

Four novel variants in *GBA1* were identified (**Table 3).** Of note, none of these were found to be present in the *GBA1*-PD browser (https://pdgenetics.shinyapps.io/gba1browser/, accessed November 1, 2024). The missense *GBA1* p.His413Arg variant (1:155235831:T:C, rs911331923, CADD=23.9, PrimateAI=0.52, AlphaMissense=0.25) was identified in a male case from the GP2-BLAAC PD cohort, with disease onset in his 40s. This variant is specific to African/African American ancestry, reported only in individuals of these ancestries in the gnomAD v4.1.0 (allele count=3) and All of Us release 7 (allele count=7) browsers, respectively. The histidine to proline change at this position was previously reported as likely pathogenic, whereas the p.His413Arg variant identified in the current study has not been reported before. The second novel missense *GBA1* p.Ile407Ser variant (1:155236249:A:C, rs1057519358, CADD=26.5, PrimateAI=0.66, AlphaMissense=0.81) was identified in a male patient of African admixed genetic ancestry from the GP2-BLAAC PD cohort with disease onset in his early 50s. While *GBA1* p.Ile407Thr was previously reported with an uncertain clinical significance, p.Ile407Ser has not been reported before. The third novel variant identified in this study was *GBA1* p.Gly364Arg (1:155236379:C:G, CADD=20.2, PrimateAI=0.69, AlphaMissense=0.22) in a male patient with slowness and/or minimal cognitive problems from the GP2-BLAAC PD dataset, who had his initial symptoms in his late 50s. While a cysteine to thymine substitution at this position was previously reported, this specific variant is novel and absent in gnomAD v4.1.0. Lastly, a frameshift variant, *GBA1* p.Ala300Profs*4 (1:155237441:GC:G, CADD=28.7), was identified in an early-onset female case of African genetic ancestry from the GP2- BLAAC PD dataset, who presented with initial PD symptoms in her early 40s and had a second-degree relative affected by the disease.

The *GBA1* p.Met400Ile variant (1:155236269:C:T, rs149487315), previously classified as having uncertain clinical significance, was the most prevalent variant among the screened PD genes, with a carrier frequency of 0.84% across all cases and an allele frequency of 0.0028 in the gnomAD African populations. This variant is more prevalent in individuals of African ancestry, with an allele count rate of 91% in the All of Us dataset (release 7) (**Supplementary Table 2**).

Although the focus of this study was not to screen for common or non-coding variants, we screened the recently identified genetic risk factor linked to PD risk in the African and African American populations (rs3115534-G). Our study identified 3,324 *GBA1-*rs3115534-G heterozygous (3,207 in African and 117 in African admixed) and 320 homozygous carriers (311 in African and nine in African admixed), with a risk allele frequency of 0.23 in cases and 0.17 in controls in African ancestry and a frequency of 0.15 in cases and 0.14 in controls in African admixed individuals. The frequency of the G allele in each dataset tested is presented in **Supplementary Table 4**.

#### The European and Asian *LRRK2* disease-causing mutational spectrum does not appear to play a major role in Parkinson’s disease in West African and African admixed populations

The current study did not identify any pathogenic or likely pathogenic *LRRK2* variants previously reported in Europeans (p.Asn1437His, p.Arg1441Ser, p.Arg1441Gly, p.Arg1441Cys, p.Arg1441His, p.Val1447Met, p.Tyr1699Cys, p.Phe1700Leu, p.Ile2020Leu, and p.Ile2020Thr) or Asian ancestries (p.Gly2385Arg). Three novel heterozygous missense *LRRK2* variants of uncertain significance were found in the All of Us and PD GENE datasets (**Table 3**). A missense p.Glu268Ala substitution (12:40243646:A:C, rs373254349, CADD=24.6, PrimateAI=0.53, AlphaMissense=0.55) was identified in a female patient with disease onset in her early 60s. A p.Ile1438Lys variant (12:40309228:A:T, CADD=24.2, PrimateAI=0.70, AlphaMissense=0.50) was found in a PD GENE participant over 60 years old (age at onset was unavailable) with a family history of PD. The third missense variant, *LRRK2* p.Arg1538Cys (12:40314047:C:T, rs150620977, CADD=29.2, PrimateAI=0.60, AlphaMissense=0.27), was detected in a female patient from All of Us, with disease onset in her early 40s and a disease duration of 15 years. The *LRRK2* p.Glu268Ala and p.Arg1538Cys variants were more prevalent in individuals of African ancestry in the All of Us dataset (release 7), with allele count rates of 93.75% African (6.25% European) and 87.5% African (8.3% American, 1.40% European, 2.78% other) respectively, across the entire dataset. The *LRRK2* p.Ile1438Lys variant was absent in both gnomAD v4.1.0 and All of Us (release 7).

#### Novel genetic variation identified in Parkinson’s disease genes warrant further replication and functional prioritization

A p.Met116Ile missense variant (4:89729236:C:A, rs1378041201, CADD=18.9, PrimateAI=0.35, AlphaMissense=0.11) in *SNCA* was identified in a male patient of African ancestry from the PD GENE dataset, with disease onset in his late 50s and no family history of PD **(Supplementary Table 2).** No clinical information about dementia was available. While a C to T transition at the same position was reported in a single individual of East Asian genetic ancestry in gnomAD, while the variant was absent in gnomAD v4.1.0.

The *VPS35* missense variant of uncertain significance p.Met607Val (16:46662991:T, rs1555523076, CADD=21.5, PrimateAI=0.81, AlphaMissense=0.18) was identified in a male PD patient of African ancestry in the NPDR Network dataset, while the p.Asp205His variant (16:46679050:C, CADD=29.4, PrimateAI=0.90, AlphaMissense=0.94) was identified in a male patient of African admixed ancestry in the GP2-BLAAC PD dataset **(Table 3)**. The individual carrying the p.Met607Val variant had disease onset in his early 40s and a positive family history, with a second-degree relative affected by the disease. At the time of enrollment, he had Hoehn and Yahr stage 2 and had demonstrated a sustained response to levodopa without dyskinesia. In terms of non-motor features, he showed no signs of hyposmia or cognitive impairment (MDS-UPDRS Part 1.1 cognition score of 0). Notably the p.Met607Val variant is 13 amino acids from the known pathogenic p.Asp620Asn variant and lies in the C terminal domain. The patient carrying the p.Asp205His variant of uncertain significance presented with initial symptoms in his late 40s and reported no family history of PD.

#### Structural variant analyses identified PRKN copy number variants linked to early-onset juvenile Parkinson’s disease in the African and African admixed populations

We screened 96 individuals for CNVs using MLPA. We prioritized samples by individuals positive for both family history and early onset PD (n=4), only family history (n=15), only early onset PD (n=36), and randomly selected sporadic PD cases (n=41). We identified one early onset PD case from the GP2- BLAAC PD dataset having a *PRKN* exon 3 heterozygous deletion and a *PRKN* exon 4 homozygous deletion (**Figure 7**). This PD case had an AAO of 27 years and no family history of PD. Mimicking the *PRKN*-PD clinical phenotype previously reported, this individual was levodopa responsive, and was positive for asymmetric onset, bradykinesia, rest tremor, rigidity, and gait difficulties. In our additional screening of 1,167 PD cases of African and African admixed ancestry using genotyping data, we identified *PRKN* CNVs in eight samples (African admixed =5, African =3), two of which were homozygous (African admixed=1, African=1). Out of these, five were early-onset cases (**Table 4, Supplementary** Figure 3). We did not identify any protein-altering or splicing *PRKN* variant among these samples. We did not identify any CNVs in *SNCA*. Further validation of the CNVs detected is needed.

#### Previously reported pathogenic variants in parkinsonism and neurodegenerative disease-related genes were not observed in African and African-admixed ancestry

We identified heterozygous variants in genes associated with recessive PD and parkinsonism, including one in *DNAJC6*, six in *VPS13C,* two in *SYNJ1*, three in *PLA2G6* and one in *FBXO7* in early-onset cases. Among these, the homozygous *FBXO7* p.Arg321Ter variant (22:32491175:C:T, rs369105683) was previously reported as pathogenic in ClinVar. However, this variant was found to be heterozygous in the current study. We also reported rare heterozygous variants in neurodegenerative disease related genes, including *TRPM7*, *MAPT*, and *APP* that could be associated with parkinsonism phenotypes; however, none have been previously reported as pathogenic. Notably, we identified a homozygous p.Ala128Gly variant (17:45982962:C:G, rs899291077) in *MAPT* in a male case from the GP2-BLAAC PD dataset. Variants in *MAPT* were previously reported in cases with progressive supranuclear palsy, an atypical parkinsonian and dementia syndrome ^35^. The carrier of this variant in our study presented with resting tremor, bradykinesia, and rigidity in his 70s, with no evidence of dementia reported (**Supplementary Table 2**). None of the cases with *APP* variants exhibited dementia or cognitive impairment. Furthermore, we identified rare variants in parkinsonism and dystonia genes, including novel *VPS13D* homozygous variants in four cases, and heterozygous variants in *PD8EB*, *KCNN2*, *CSF1R*, *THAP1*, *TOR1A*, *ANO3, GCH1, VPS16, KCDT17, KMT2B*, and *ATP1A3* genes. These variants were novel or previously reported with unknown clinical significance (**Supplementary Table 5**).

#### Runs of homozygosity analysis highlights regions overlapping known Parkinson’s disease, pallido-pyramidal syndrome, and atypical parkinsonism gene regions and risk loci

In the African and African admixed groups, 164 and 43 ROH pools, respectively, were identified, which overlap known PD and atypical parkinsonism gene regions and GWAS loci, respectively (**Supplementary Tables 6 and 7)**. None of the ROH regions enriched in cases for either group passed the Bonferroni correction. However, in the African group, one ROH pool (S2421) that overlaps the *BAG3* and *INPP5F* genes was present in four cases and completely absent in controls. Moreover, in the African admixed group, two ROH pools (S622 and S636) were found in cases and absent in controls. These pools overlap the *MEX3C* (n=2 cases), *SMAD4* (n=2 cases), and *PLA2G6* (n=2 cases) genes (**Supplementary Tables 6 and 7)**.

## Discussion

Here we undertook the largest sequencing characterization of PD, parkinsonism, and neurodegenerative diseases related genes in the African and African admixed populations. We created the most comprehensive genetic catalog of both known and novel coding and splicing variants potentially linked to PD etiology in these populations.

Biallelic *GBA1* variants are associated with Gaucher’s disease, while monoallelic *GBA1* variants are recognized as the most prevalent genetic risk factor for PD, with their prevalence varying among diverse ethnic groups ^36–38,39–41^. In this study, we identified that heterozygous rare and potentially disease-causing *GBA1* variants represent the most commonly identified mutations in PD among patients of African and African admixed ancestry, with a frequency of 4%, which aligns with the frequency of 4.6% reported in North African populations ^42^. Our study identified 18 heterozygous protein-altering variants in *GBA1*, five of which seem to be more frequent in African ancestry than other ancestry populations according to gnomAD v4.1 and All of Us (release 7), including p.His413Arg, p.Met400Ile, p.Asn227Ser, p.Ser77Arg, and p.Thr75del. Of those, p.Asn227Ser was classified as severe, p.Thr75del as mild, p.Met400Ile as unknown, while p.His413Arg and p.Ser77Arg variants have not been previously reported ^43^. Of note, no carriers were identified for the most common disease-causing *GBA1* variants in the Ashkenazi Jewish and European populations (p.Asn409Ser, p.Leu483Pro, p.Thr408Met and p.Glu365Lys).

Recently, the first PD GWAS in the African and African admixed ancestry populations revealed a novel non-coding and common variant within *GBA1* (rs3115534-G) conferring a major population attributable risk to disease in the patient population ^6^. Follow-up functional characterization highlighted a novel mechanism of *GBA1*-related PD risk, offering a promising target for precision medicine in a population that has been historically underserved in genetic research and therapeutic development ^7^. Though this study was primarily focused on assessing the impact of coding variants, we specifically screened for this variant in our cohort. Our analyses showed a higher estimate of the risk G allele in predicted African ancestry individuals, with an overall frequency of 0.23 in cases and 0.17 in controls and a frequency of 0.15 in cases and 0.14 in controls among individuals of African admixed ancestry.

Our results showed that the previously reported European and Asian *LRRK2* disease-causing mutation spectrum does not seem to have a significant role in PD in West African and African American populations. For instance, the *LRRK2* p.Gly2019Ser variant was not present in any of the tested cases. It is known that *LRRK2* p.Gly2019Ser shows varying frequencies across North African genetic ancestry groups, ranging from 10-42%, with the highest rates reported in Tunisia ^44–50^. Conversely, this variant is rare among Sub-Saharan Africans, with only a few cases reported in South Africa ^51^. A recent study screening for *LRRK2* p.Gly2019Ser in 109 Nigerian patients did not identify any variant carriers ^52^. Additionally, *LRRK2* p.Gly2019Ser carriers were not detected among 16 South African patients ^53^ and 38 Zambian individuals with PD ^54^. Our results are in concordance with a recent study examining *LRRK2* mutations in 22 African American PD patients ^3^. Although the authors identified both known and novel variants, no previously reported pathogenic mutations were detected, in contrast to what has been observed in North African populations. Interestingly, we identified a heterozygous carrier of the *LRRK2* p.Gly8Arg variant, a novel coding mutation not previously reported in the literature, considered to be of uncertain significance, and only present in populations of African and Admixed American ancestry according to gnomAD v4.1.0 and All of Us browsers (release 7). Similarly, the *LRRK2* p.Glu1797Asp heterozygous variant was identified in our study among two male PD cases of African ancestry; this variant has been shown to be present in African and Admixed American ancestry populations and absent in Europeans (73% African and 23% Admixed American ancestry in All of Us release 7). Overall, these findings highlight ancestral differences in *LRRK2* variant distribution, underscoring the need for further research to explore these disparities. The discovery of novel variants opens new avenues for future functional investigations.

Our study aligns with previous research showing that *SNCA* coding and structural variation is rare in populations of African ancestry ^55^. In fact, our analysis did not detect any coding variants or structural rearrangements in *SNCA* across the cohorts studied. This finding is consistent with several other studies on PD in African and African admixed populations. For example, a study of 38 individuals with PD from Zambia (plus one with atypical parkinsonism) found no pathogenic *SNCA* mutations ^54^. Similarly, a screening of 15 early-onset PD cases from Southwestern Nigeria ^52^, and a study of 202 South African individuals with PD (8% Black) also found no exonic *SNCA* variants linked to disease ^56^.

Regarding recessive genes, nearly 20 PD cases associated with *PRKN* and *PINK1* mutations have been documented in Sub-Saharan Africa across multiple studies ^54,55,57–61^. In contrast, fewer than 10 PD cases with these mutations have been reported in North Africa ^50,62–67^. Though compound heterozygous *PRKN* mutations cause recessive disease, heterozygous protein altering variants do not appear to confer PD risk, with multiple null associations reported ^68–70^. In our study, we identified two heterozygous *PRKN* variants in an early-onset PD case, one of which had been previously reported as pathogenic, suggesting a potential compound heterozygous state that was not confirmed due to the lack of phasing data. Together with the CNVs identified in eight cases—three from African and five from African admixed ancestry —a total of 14 early-onset PD cases were found to carry potential disease-causing *PRKN* mutations (**Table 2, <u>Table3</u>, Supplementary Table 2**).

*PRKN* carriers constituted 1.3% (9 out of 710) of all cases. *PRKN* variants constituted 13.6% of all observed variants in PD genes, with 13 variants in the GP2-BLAAC PD cohort and 10 in the NPDR Network. Of those, 9 variants were more frequent in cases than controls and 7 were more frequent in African ancestry individuals, relative to non-Finnish European ancestry, according to gnomAD. We identified two heterozygous *PINK1* variants, previously reported as pathogenic, in two late-onset PD cases, which were likely not the cause of underlying PD in these cases. While we did not report any pathogenic variants in *DJ-1* and *VPS13C*, we found four novel *DJ-1* and 13 novel *VPS13C* variants, all in heterozygous state, that warrant further study.

Several limitations should be acknowledged. First, our study includes a highly heterogeneous population. The current available ancestry reference panels cannot accurately distinguish between African subpopulations. African admixed study participants commonly exhibit a diverse heritage often encompassing African, European, and, in certain instances, Native American genetic components. Although reported proportions vary, the predominant ancestry of African Americans is typically traced back to Niger-Kordofanian (∼71-73%), European (∼13-24%), and other African (∼0.8-8%) populations ^71,72^. The African component of African American ancestry is itself varied, reflecting the multitude of ethnic groups involved in the trans-Atlantic slave trade. It is important to note that most of the PD studies encompassing individuals of African ancestry primarily stem from West African populations, and while they capture a portion of the genetic diversity present in Africa, they do not represent the continent in its entirety ^71^. Second, the majority of the candidate variants identified in our study are singletons, observed only in a single individual or within a single dataset. Therefore, caution is warranted in interpreting their causality. Future studies should aim to validate these findings in larger datasets or functional assays to better understand their potential role in disease. In addition, the variants across the *GBA1* and *GBAP1* homology region require further analysis for validation using other tools, since Gauchian is unable to detect novel variants in the *GBA1* and *GBAP1* homology region. Furthermore, certain potentially impactful variants might have been missed, particularly in regions of the genome that are difficult to resolve with short-read sequencing approaches. Long-read technologies are more effective in detecting large insertions or deletions, which may not be captured by short-read sequencing or MLPA. Incorporating long-read sequencing in future studies could enhance our understanding of the genetic underpinnings of PD. Clinical information was also lacking as only a subset of samples had data from the Montreal Cognitive Assessment (MoCA) and limited data was collected for the Identification and Intervention for Dementia in Elderly Africans (IDEA) cognitive screen and Rapid Eye Movement sleep behavior disorder (RBD). One question from the CISI-PD questionnaire was used to assess cognitive status. Future clinical data collection efforts as part of the GP2-BLAAC PD study will include the MoCA and other clinical assessments.

Our study, the largest of its kind to explore rare coding and splicing variations in African and African admixed populations, aims to fill critical gaps in PD genetics research by providing a comprehensive genetic characterization of these ancestries. While there is growing recognition of the importance of studying PD in diverse populations, Black and African admixed communities have historically been underrepresented in research, resulting in significant gaps in understanding how PD manifests and progresses. By focusing on these underserved groups, our work seeks to enhance our current knowledge of PD etiology, ultimately improving diagnostic accuracy and therapeutic development. As we advance toward precision medicine, therapies should be developed with a global perspective. Operating under the umbrella of the GP2, it is essential that the data used to guide individualized treatments reflects the diversity of all populations, ensuring equitable benefits from medical advancements in PD care.

## Funding

This work utilized the computational resources of the NIH HPC Biowulf cluster (https://hpc.nih.gov). This research was supported in part by the Intramural Research Program of the NIH, National Institute on Aging (NIA), National Institutes of Health, Department of Health and Human Services; project number ZO1 AG000535 and ZIA AG000949, as well as the National Institute of Neurological Disorders and Stroke (NINDS, program # ZIANS003154) and the National Human Genome Research Institute (NHGRI). This work utilized the computational resources of the NIH STRIDES Initiative (https://cloud.nih.gov) through the Other Transaction agreement - Azure: OT2OD032100, Google Cloud Platform: OT2OD027060, Amazon Web Services: OT2OD027852. Data used in the preparation of this article were obtained from the Global Parkinson’s Genetics Program (GP2). GP2 is funded by the Aligning Science Across Parkinson’s (ASAP) initiative and implemented by The Michael J. Fox Foundation for Parkinson’s Research (https://gp2.org). For a complete list of GP2 members see https://gp2.org. Additional funding was provided by The Michael J. Fox Foundation for Parkinson’s Research through grant MJFF-009421/17483.

## Acknowledgements

We thank Paige Brown Jarreau for her meticulous editing of this manuscript.

Data used in the preparation of this article were obtained from Global Parkinson’s Genetics Program (GP2). GP2 is funded by the Aligning Science Across Parkinson’s (ASAP) initiative and implemented by The Michael J. Fox Foundation for Parkinson’s Research (https://gp2.org). For a complete list of GP2 members see https://gp2.org. The All of Us Research Program is supported by the National Institutes of Health, Office of the Director: Regional Medical Centers: 1 OT2 OD026549; 1 OT2 OD026554; 1 OT2 OD026557; 1 OT2 OD026556; 1 OT2 OD026550; 1 OT2 OD 026552; 1 OT2 OD026553; 1 OT2 OD026548; 1 OT2 OD026551; 1 OT2 OD026555; IAA #: AOD 16037; Federally Qualified Health Centers: HHSN 263201600085U; Data and Research Center: 5 U2C OD023196; Biobank: 1 U24 OD023121; The Participant Center: U24 OD023176; Participant Technology Systems Center: 1 U24 OD023163; Communications and Engagement: 3 OT2 OD023205; 3 OT2 OD023206; and Community Partners: 1 OT2 OD025277; 3 OT2 OD025315; 1 OT2 OD025337; 1 OT2 OD025276. In addition, the All of Us Research Program would not be possible without the partnership of its participants.

This research has been conducted using the UK Biobank Resource under application number 33601.

## Competing interests

KL, DV, HLL, HI, MJK and MAN declare that they are consultants employed by DataTecnica LLC, whose participation in this is part of a consulting agreement between the US National Institutes of Health and said company. MAN also owns stock from Neuron23 Inc and Character Biosciences.

## Data availability

All GP2 data for these analyses is available through collaboration with Accelerating Medicines Partnership in Parkinson’s disease and is available through application on the AMP-PD website (https://amp-pd.org/register-for-amp-pd). Other publicly available data consortiums include All of Us Research Program (https://www.researchallofus.org/register/) and UK Biobank (https://www.ukbiobank.ac.uk/enable-your-research/register), in which data can be accessed after applying. We have received an exception to the Data and Statistics Dissemination Policy from the All of Us Resource Access Board.

## Code availability

Analyses such as genotyping quality control, ancestry prediction, and processing were performed using GenoTools (https://github.com/dvitale199/GenoTools). A repository of all other code used for processing and analyzing is publicly available (https://github.com/GP2code/PD-GeneticCharacterization-inAFRandAAC/; DOI 10.5281/zenodo.14579574).

## Authors roles

SBC, NO, CB and AS contributed to the conception and design of the study.

FA, PWC, SG, PS, MK, PL, KP, RR, and SBC contributed to the acquisition and analysis of the data. FA, KP, PWC, PS, and SBC drafted significant portions of the manuscript or figures.

All the authors contributed to editing and critically reviewing the manuscript

---

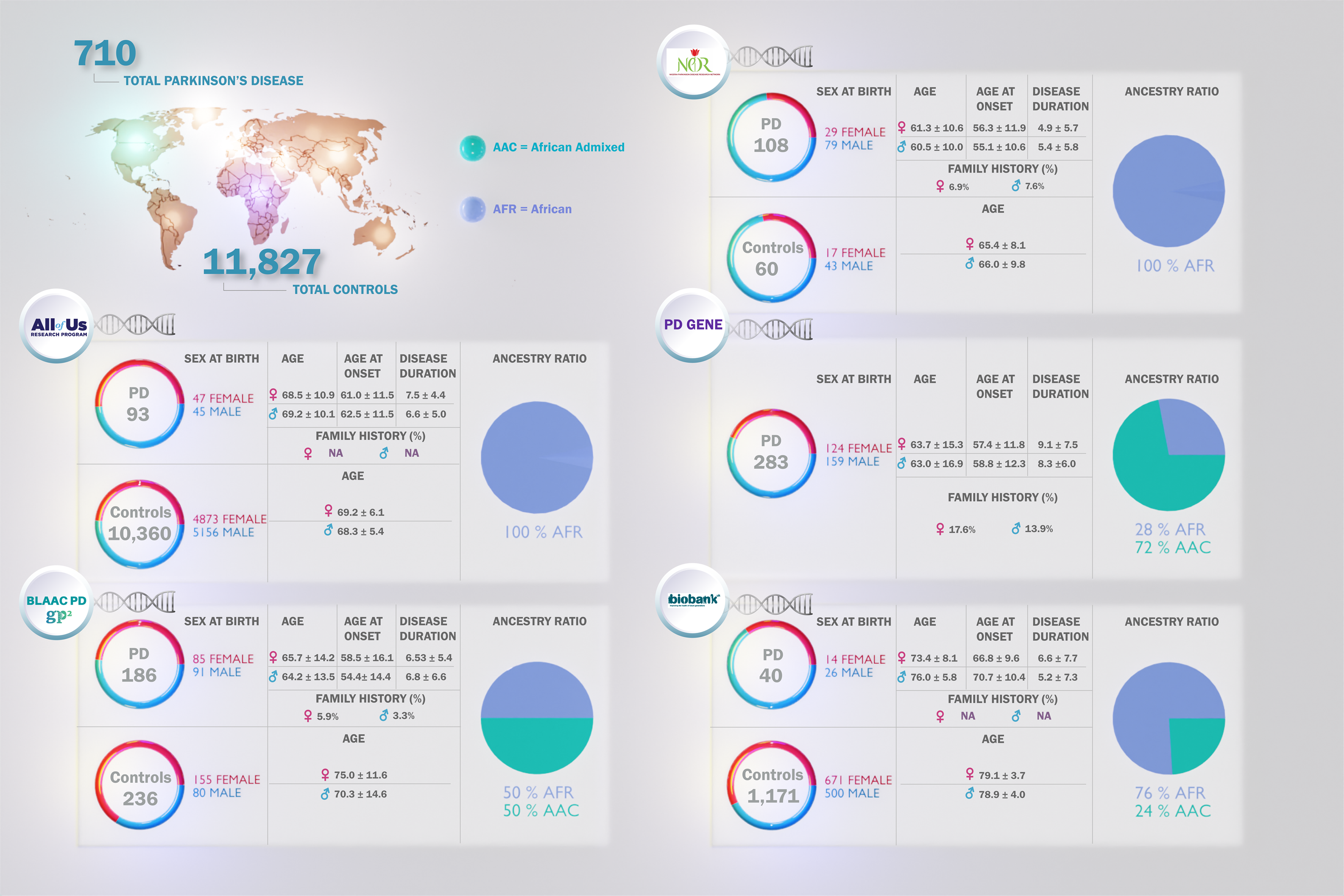

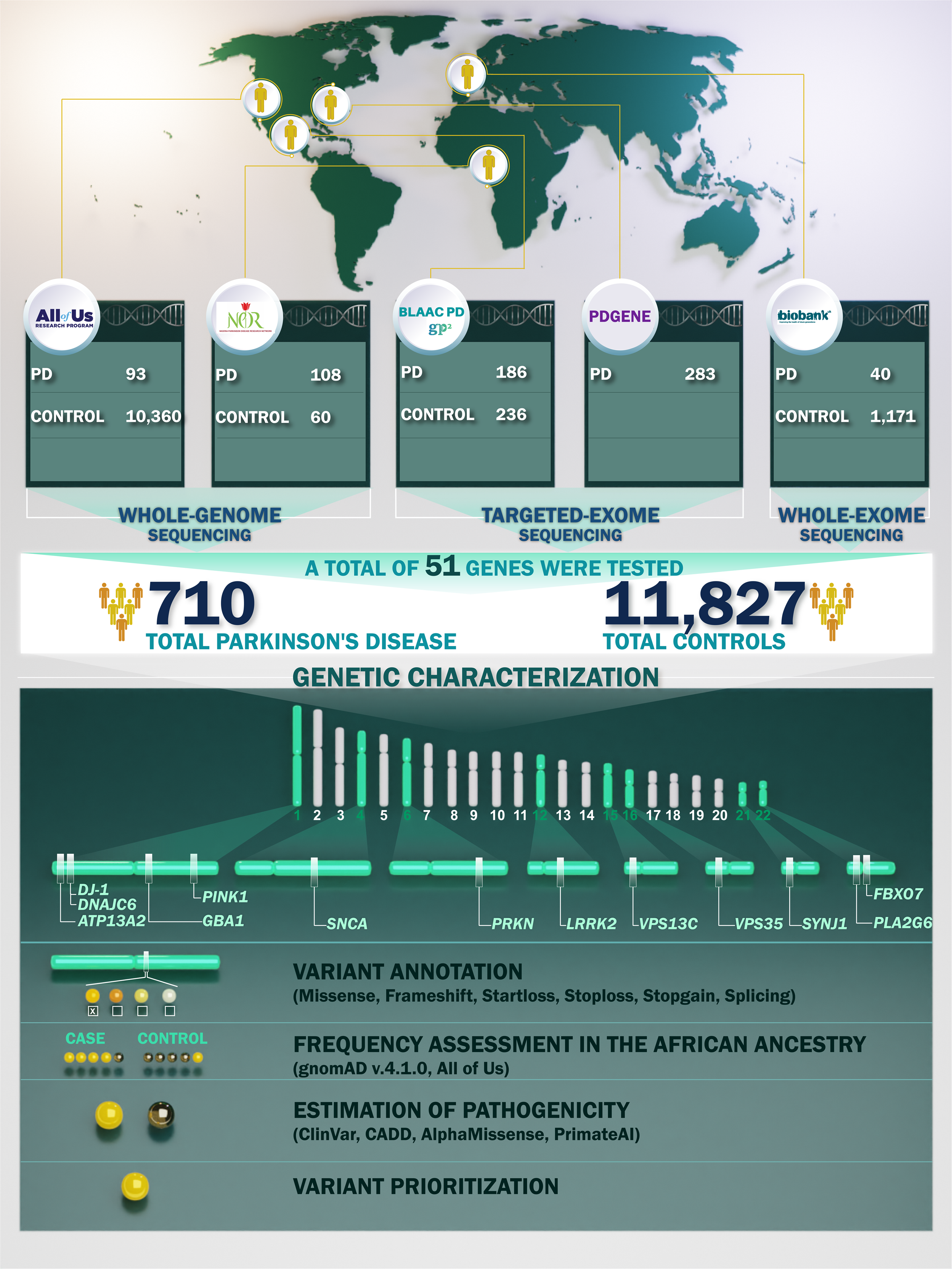

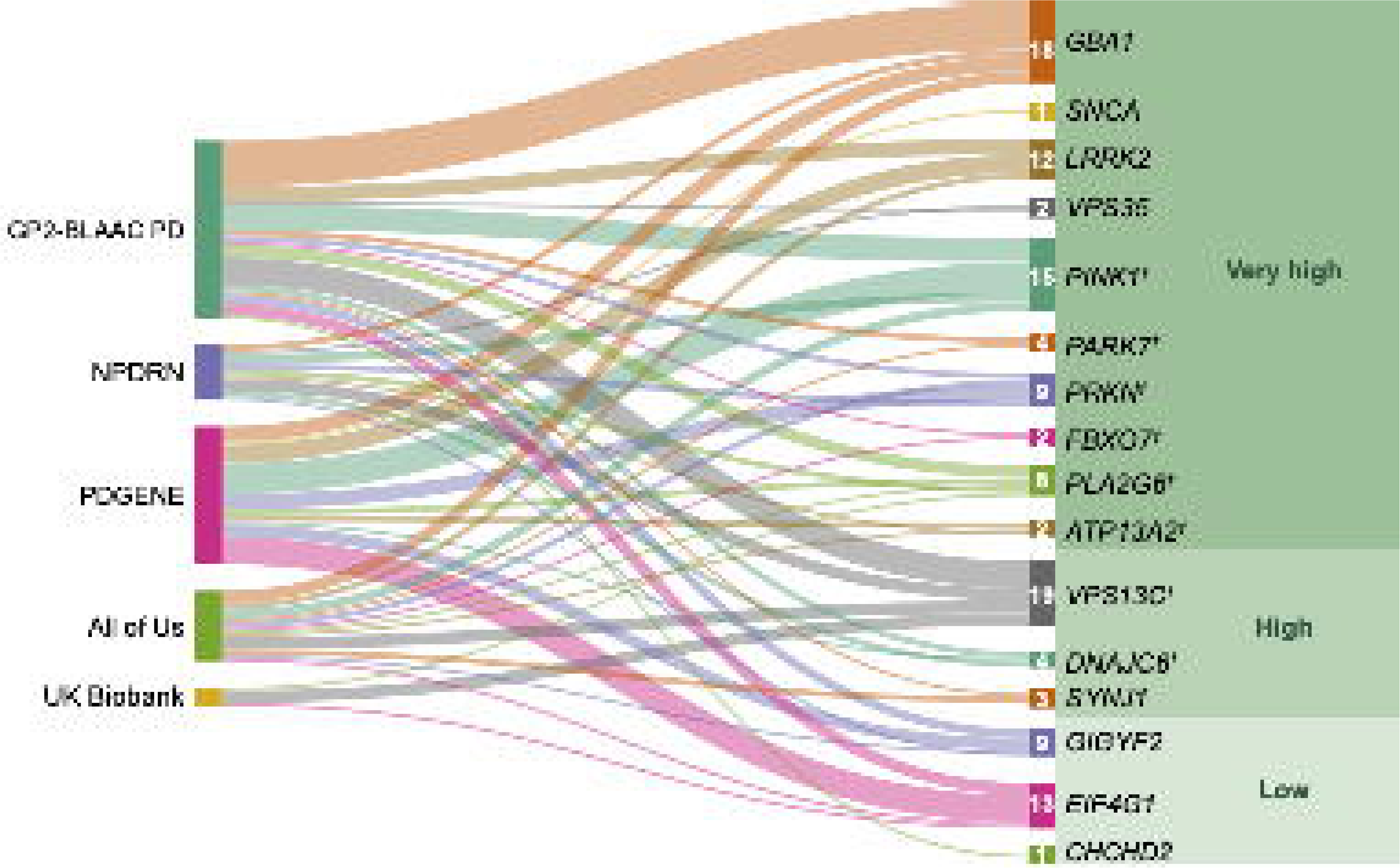

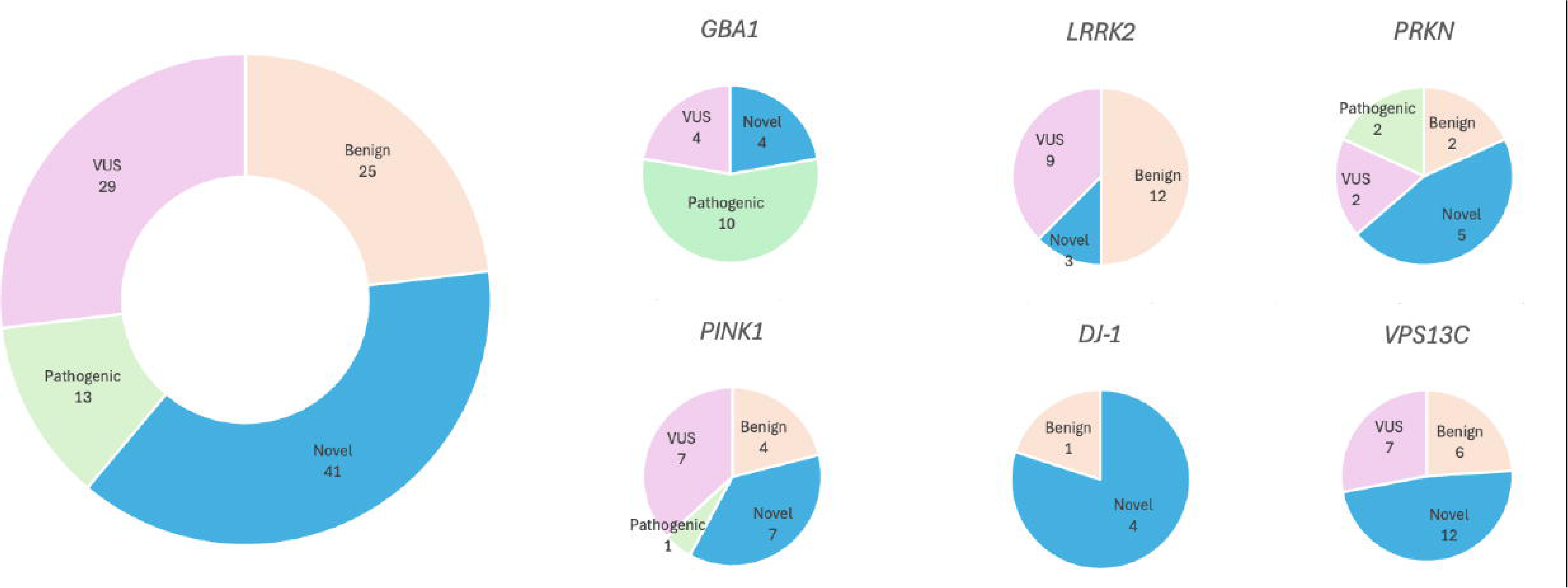

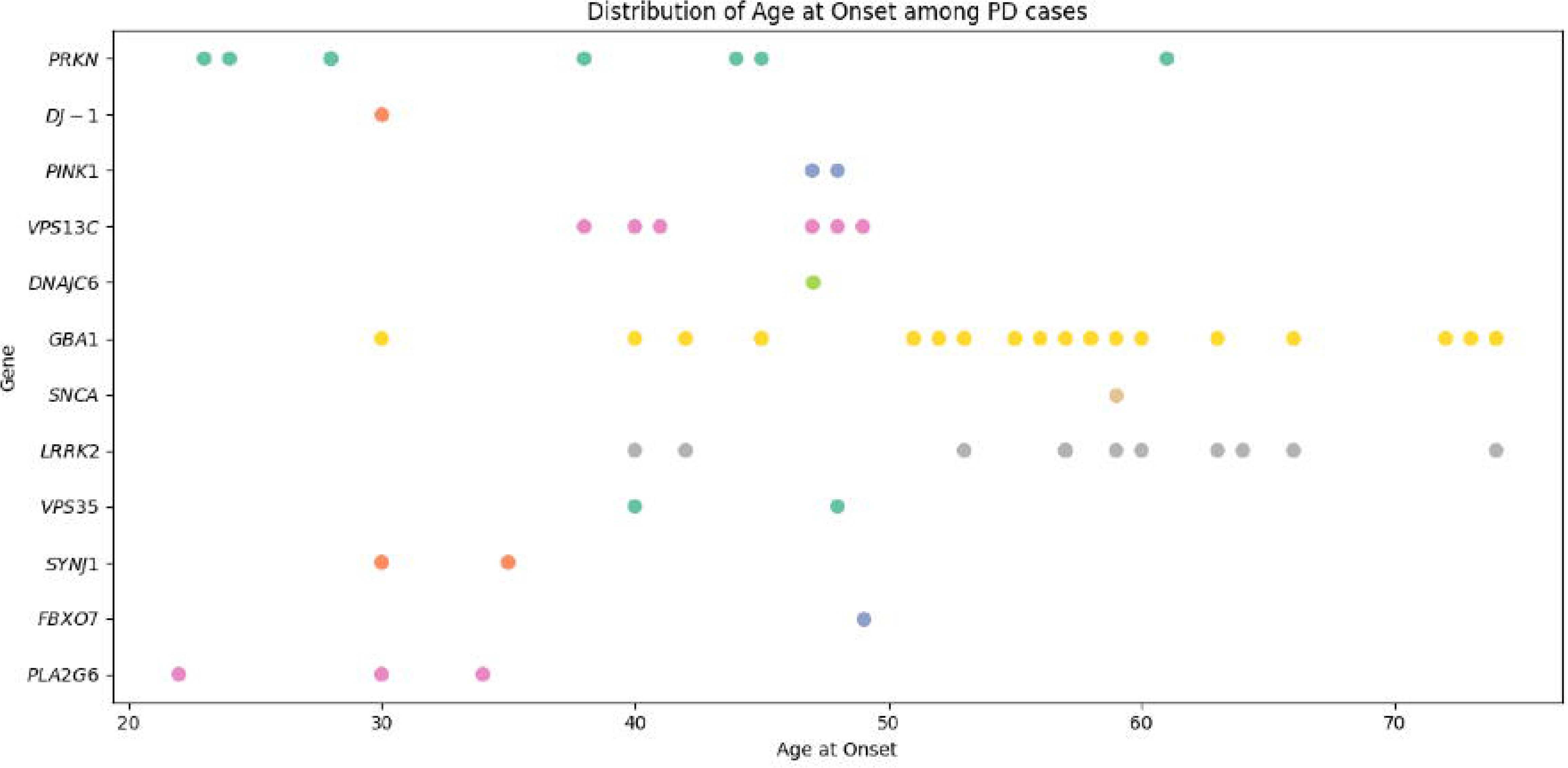

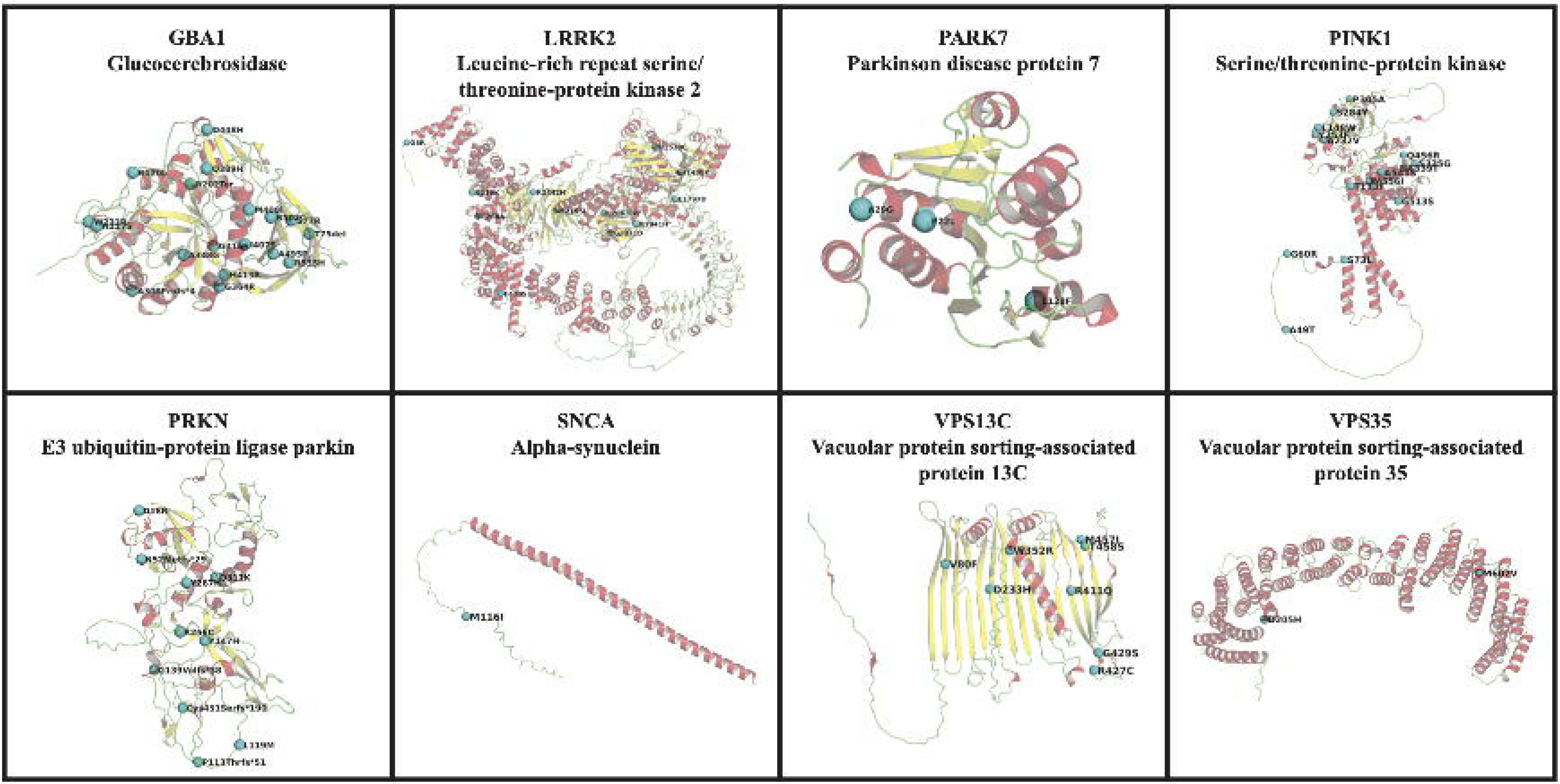

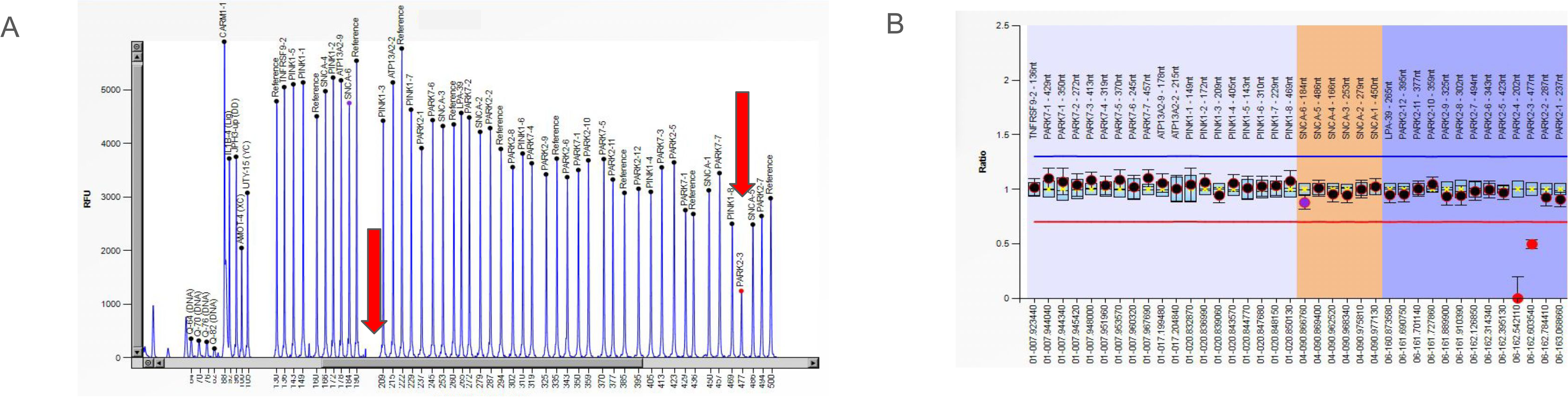

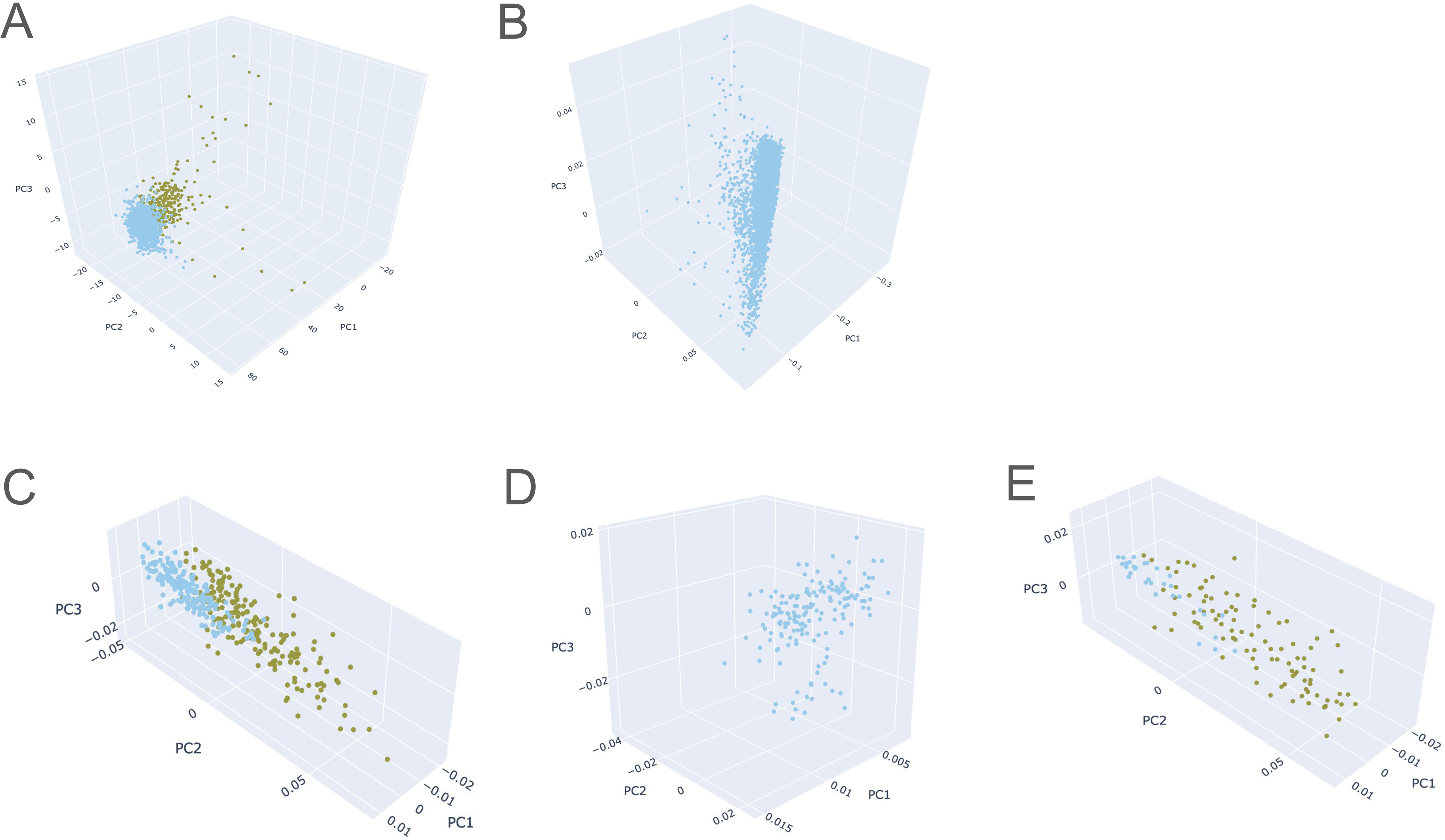

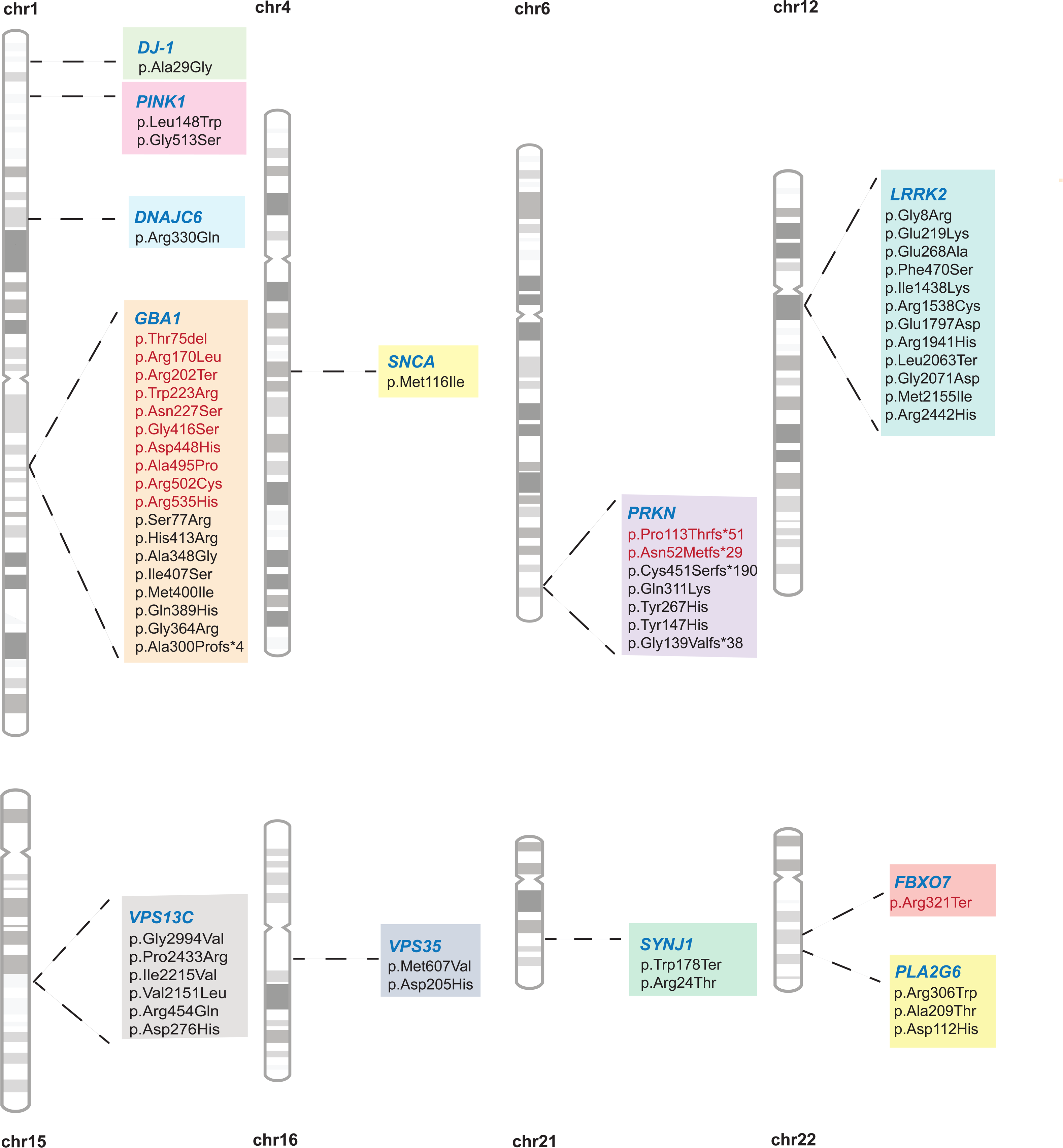

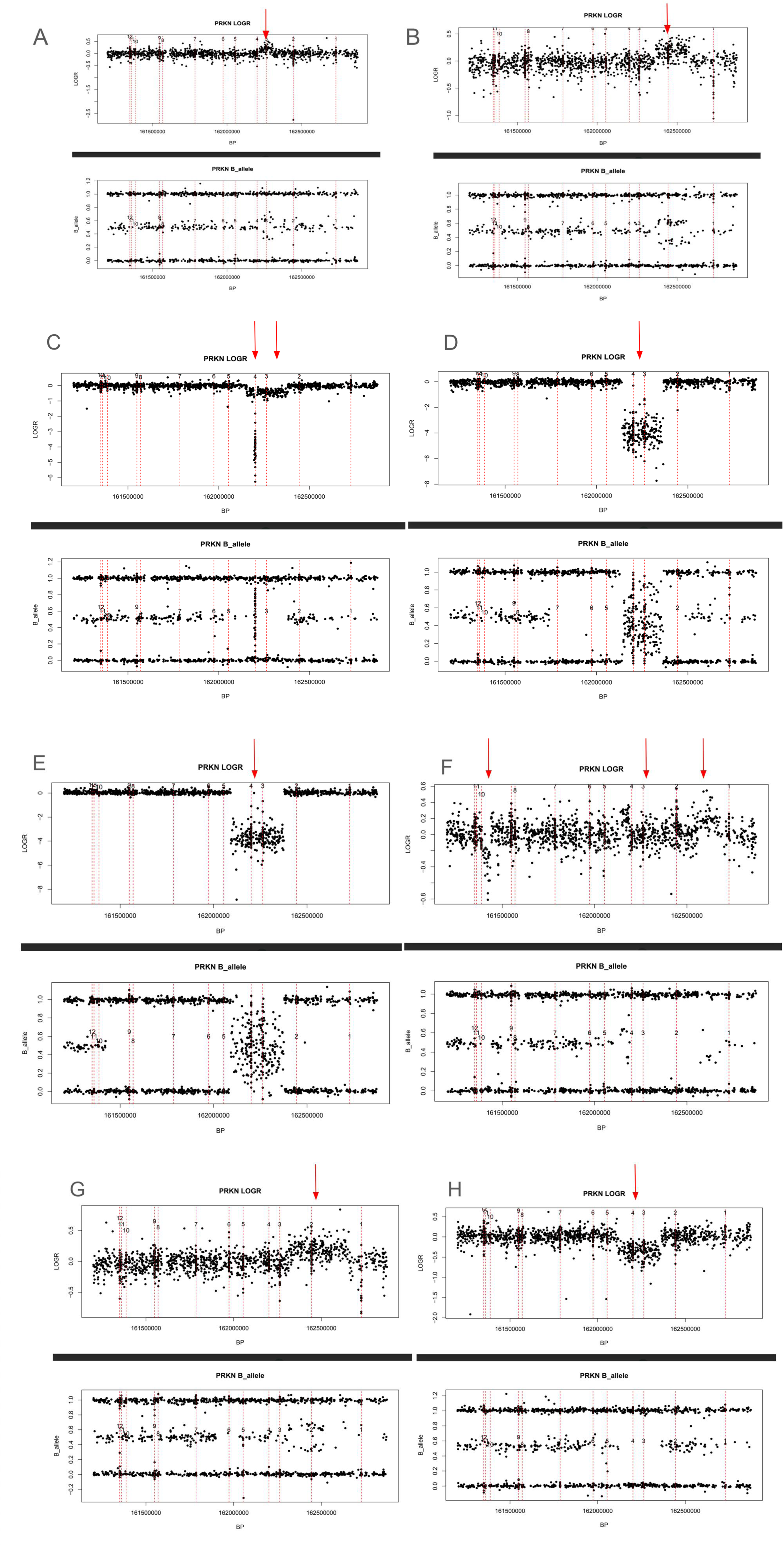

## Notes

### Clinical Protocols

https://github.com/GP2code/PD-GeneticCharacterization-inAFRandAAC/

### Author Declarations

All GP2 data for these analyses is available through collaboration with Accelerating Medicines Partnership in Parkinson′s disease and is available through application on the AMP—PD website. Other publicly available data consortiums include All of Us Research Program and UK Biobank, in which data can be accessed after applying. We have received an exception to the Data and Statistics Dissemination Policy from the All of Us Resource Access Board.

### Summary of Updates

Some typological errors of the author names and their affiliations were updated. Supplementary table 4 was updated, to remove the frequency of the Nigerian cohort from the total frequency estimation. The text related to Supplementary Table 4 was revised accordingly.

## References

1. Khani M, Cerquera-Cleves C, Kekenadze M, Wild Crea P, Singleton AB, Bandres-Ciga S. Towards a global view of Parkinson’s disease genetics. Ann Neurol. 2024;95(5):831–842.

2. Okunoye O, Zewde YZ, Azar J, et al. The State of Play of Parkinson’s Disease in Africa: A Systematic Review and Point of View. medRxiv. Published online July 8, 2023:2023.07.07.23292392. doi:10.1101/2023.07.07.23292392

3. Ross OA, Wilhoite GJ, Bacon JA, et al. LRRK2 variation and Parkinson’s disease in African Americans. Mov Disord. 2010;25(12):1973–1976.

4. McGuire V, Van Den Eeden SK, Tanner CM, et al. Association of DRD2 and DRD3 polymorphisms with Parkinson’s disease in a multiethnic consortium. J Neurol Sci. 2011;307(1-2):22–29.

5. Clark LN, Levy G, Tang MX, et al. The Saitohin “Q7R” polymorphism and tau haplotype in multi-ethnic Alzheimer disease and Parkinson’s disease cohorts. Neurosci Lett. 2003;347(1):17–20.

6. Rizig M, Bandres-Ciga S, Makarious MB, et al. Identification of genetic risk loci and causal insights associated with Parkinson’s disease in African and African admixed populations: a genome-wide association study. Lancet Neurol. 2023;22(11):1015–1025.

7. Álvarez Jerez P, Wild Crea P, Ramos DM, et al. African ancestry neurodegeneration risk variant disrupts an intronic branchpoint in GBA1. Nature Structural & Molecular Biology. Published online December 12, 2024:1–9.

8. Mahungu AC, Anderson DG, Rossouw AC, et al. Screening of the glucocerebrosidase (GBA) gene in South Africans of African ancestry with Parkinson’s disease. Neurobiol Aging. 2020;88:156.e11–e156.e14.

9. Milanowski LM, Oshinaike O, Walton RL, et al. Screening of GBA Mutations in Nigerian Patients with Parkinson’s Disease. Mov Disord. 2021;36(12):2971–2973.

10. Bandres-Ciga S, Black and African American Connections to Parkinson’s Disease (BLAAC PD) Study Group. Black and African American Connections to Parkinson’s Disease Study: Addressing Missing Diversity in Parkinson’s Disease Genetics. Mov Disord. 2022;37(7):1559–1561.

11. Global Parkinson’s Genetics Program. GP2: The global Parkinson’s genetics program. Mov Disord. 2021;36(4):842–851.

12. Ojo OO, Abubakar SA, Iwuozo EU, et al. The Nigeria Parkinson Disease Registry: Process, Profile, and Prospects of a Collaborative Project. Mov Disord. 2020;35(8):1315–1322.

13. Cook L, Verbrugge J, Schwantes-An TH, et al. Parkinson’s disease variant detection and disclosure: PD GENEration, a North American study. Brain. 2024;147(8):2668–2679.

14. All of Us Research Program Investigators, Denny JC, Rutter JL, et al. The “All of Us” Research Program. N Engl J Med. 2019;381(7):668–676.

15. Bycroft C, Freeman C, Petkova D, et al. The UK Biobank resource with deep phenotyping and genomic data. Nature. 2018;562(7726):203-209.

16. Westenberger A, Skrahina V, Usnich T, et al. Relevance of genetic testing in the gene-targeted trial era: the Rostock Parkinson’s disease study. Brain. 2024;147(8):2652–2667.

17. Chahine LM, Louie N, Solle J, et al. The Black and African American Connections to Parkinson’s Disease (BLAAC PD) study protocol. BMC Neurol. 2024;24(1):403.

18. Martínez-Martín P, Forjaz MJ, Cubo E, Frades B, de Pedro Cuesta J, ELEP Project Members. Global versus factor-related impression of severity in Parkinson’s disease: a new clinimetric index (CISI-PD). Mov Disord. 2006;21(2):208–214.

19. Postuma RB, Berg D, Stern M, et al. MDS clinical diagnostic criteria for Parkinson’s disease: MDS-PD Clinical Diagnostic Criteria. Mov Disord. 2015;30(12):1591–1601.

20. Hughes AJ, Daniel SE, Kilford L, Lees AJ. Accuracy of clinical diagnosis of idiopathic Parkinson’s disease: a clinico-pathological study of 100 cases. J Neurol Neurosurg Psychiatry. 1992;55(3):181–184.

21. Manichaikul A, Mychaleckyj JC, Rich SS, Daly K, Sale M, Chen WM. Robust relationship inference in genome-wide association studies. Bioinformatics. 2010;26(22):2867–2873.

22. Purcell S, Neale B, Todd-Brown K, et al. PLINK: a tool set for whole-genome association and population-based linkage analyses. Am J Hum Genet. 2007;81(3):559–575.

23. Simón-Sánchez J, Kilarski LL, Nalls MA, et al. Cooperative genome-wide analysis shows increased homozygosity in early onset Parkinson’s disease. PLoS One. 2012;7(3):e28787.

24. Nalls MA, Blauwendraat C, Vallerga CL, et al. Identification of novel risk loci, causal insights, and heritable risk for Parkinson’s disease: a meta-analysis of genome-wide association studies. Lancet Neurol. 2019;18(12):1091–1102.

25. Kim JJ, Vitale D, Otani DV, et al. Multi-ancestry genome-wide association meta-analysis of Parkinson’s disease. Nat Genet. 2024;56(1):27–36.

26. Loesch DP, Horimoto ARVR, Heilbron K, et al. Characterizing the Genetic Architecture of Parkinson’s Disease in Latinos. Ann Neurol. 2021;90(3):353–365.

27. Foo JN, Chew EGY, Chung SJ, et al. Identification of Risk Loci for Parkinson Disease in Asians and Comparison of Risk Between Asians and Europeans: A Genome-Wide Association Study. JAMA Neurol. 2020;77(6):746–754.

28. Bandrés-Ciga S, Price TR, Barrero FJ, et al. Genome-wide assessment of Parkinson’s disease in a Southern Spanish population. Neurobiol Aging. 2016;45:213.e3–e213.e9.

29. Wang K, Li M, Hakonarson H. ANNOVAR: functional annotation of genetic variants from high-throughput sequencing data. Nucleic Acids Res. 2010;38(16):e164.

30. Toffoli M, Chen X, Sedlazeck FJ, et al. Comprehensive short and long read sequencing analysis for the Gaucher and Parkinson’s disease-associated GBA gene. Commun Biol. 2022;5(1):670.

31. Kircher M, Witten DM, Jain P, O’Roak BJ, Cooper GM, Shendure J. A general framework for estimating the relative pathogenicity of human genetic variants. Nat Genet. 2014;46(3):310–315.

32. Blauwendraat C, Nalls MA, Singleton AB. The genetic architecture of Parkinson’s disease. Lancet Neurol. 2020;19(2):170–178.

33. Jumper J, Evans R, Pritzel A, et al. Highly accurate protein structure prediction with AlphaFold. Nature. 2021;596(7873):583-589.

34. Varadi M, Bertoni D, Magana P, et al. AlphaFold Protein Structure Database in 2024: providing structure coverage for over 214 million protein sequences. Nucleic Acids Res. 2024;52(D1):D368–D375.

35. Hutton M, Lendon CL, Rizzu P, et al. Association of missense and 5’-splice-site mutations in tau with the inherited dementia FTDP-17. Nature. 1998;393(6686):702-705.

36. Migdalska-Richards A, Schapira AHV. The relationship between glucocerebrosidase mutations and Parkinson disease. J Neurochem. 2016;139 Suppl 1(Suppl Suppl 1):77–90.

37. De Marco EV, Annesi G, Tarantino P, et al. Glucocerebrosidase gene mutations are associated with Parkinson’s disease in southern Italy. Mov Disord. 2008;23(3):460–463.

38. Kalinderi K, Bostantjopoulou S, Paisan-Ruiz C, Katsarou Z, Hardy J, Fidani L. Complete screening for glucocerebrosidase mutations in Parkinson disease patients from Greece. Neurosci Lett. 2009;452(2):87–89.

39. Tan EK, Tong J, Fook-Chong S, et al. Glucocerebrosidase mutations and risk of Parkinson disease in Chinese patients. Arch Neurol. 2007;64(7):1056–1058.

40. Mitsui J, Mizuta I, Toyoda A, et al. Mutations for Gaucher disease confer high susceptibility to Parkinson disease. Arch Neurol. 2009;66(5):571–576.

41. Velez-Pardo C, Lorenzo-Betancor O, Jimenez-Del-Rio M, et al. The distribution and risk effect of GBA variants in a large cohort of PD patients from Colombia and Peru. Parkinsonism Relat Disord. 2019;63:204–208.

42. Lesage S, Condroyer C, Hecham N, et al. Mutations in the glucocerebrosidase gene confer a risk for Parkinson disease in North Africa. Neurology. 2011;76(3):301–303.

43. Parlar SC, Grenn FP, Kim JJ, Baluwendraat C, Gan-Or Z. Classification of GBA1 Variants in Parkinson’s Disease: The GBA1-PD Browser. Mov Disord. 2023;38(3):489–495.

44. Belarbi S, Hecham N, Lesage S, et al. LRRK2 G2019S mutation in Parkinson’s disease: A neuropsychological and neuropsychiatric study in a large Algerian cohort. Parkinsonism Relat Disord. 2010;16:676–679.

45. Troiano AR, Elbaz A, Lohmann E, et al. Low disease risk in relatives of north african lrrk2 Parkinson disease patients. Neurology. 2010;75(12):1118–1119.

46. Hashad DI, Abou-Zeid AA, Achmawy GA, Allah HMOS, Saad MA. G2019S mutation of the leucine-rich repeat kinase 2 gene in a cohort of Egyptian patients with Parkinson’s disease. Genet Test Mol Biomarkers. 2011;15(12):861–866.

47. Warren L, Gibson R, Ishihara L, et al. A founding LRRK2 haplotype shared by Tunisian, US, European and Middle Eastern families with Parkinson’s disease. Parkinsonism Relat Disord. 2008;14:77–80.

48. Ishihara L, Gibson RA, Warren L, et al. Screening for Lrrk2 G2019S and clinical comparison of Tunisian and North American Caucasian Parkinson’s disease families. Mov Disord. 2007;22(1):55–61.

49. Hulihan MM, Ishihara-Paul L, Kachergus J, et al. LRRK2 Gly2019Ser penetrance in Arab-Berber patients from Tunisia: a case-control genetic study. Lancet Neurol. 2008;7(7):591–594.

50. Nishioka K, Kefi M, Jasinska-Myga B, et al. A comparative study of LRRK2, PINK1 and genetically undefined familial Parkinson’s disease. J Neurol Neurosurg Psychiatry. 2010;81(4):391–395.

51. du Toit N, van Coller R, Anderson DG, Carr J, Bardien S. Frequency of the LRRK2 G2019S mutation in South African patients with Parkinson’s disease. Neurogenetics. 2019;20(4):215–218.

52. Milanowski LM, Oshinaike O, Broadway BJ, et al. Early-Onset Parkinson Disease Screening in Patients From Nigeria. Front Neurol. 2020;11:594927.

53. Mahne AC, Carr JA, Bardien S, Schutte CM. Clinical findings and genetic screening for copy number variation mutations in a cohort of South African patients with Parkinson’s disease. S Afr Med J. 2016;106(6). doi:10.7196/SAMJ.2016.v106i6.10340

54. Yonova-Doing E, Atadzhanov M, Quadri M, et al. Analysis of LRRK2, SNCA, Parkin, PINK1, and DJ-1 in Zambian patients with Parkinson’s disease. Parkinsonism Relat Disord. 2012;18(5):567–571.

55. Keyser RJ, Lombard D, Veikondis R, Carr J, Bardien S. Analysis of exon dosage using MLPA in South African Parkinson’s disease patients. Neurogenetics. 2010;11(3):305–312.

56. Keyser RJ, Oppon E, Carr JA, Bardien S. Identification of Parkinson’s disease candidate genes using CAESAR and screening of MAPT and SNCAIP in South African Parkinson’s disease patients. J Neural Transm (Vienna). 2011;118(6):889–897.

57. Bardien S, Keyser R, Yako Y, Lombard D, Carr J. Molecular analysis of the parkin gene in South African patients diagnosed with Parkinson’s disease. Parkinsonism Relat Disord. 2009;15(2):116–121.

58. Haylett WL, Keyser RJ, du Plessis MC, et al. Mutations in the parkin gene are a minor cause of Parkinson’s disease in the South African population. Parkinsonism Relat Disord. 2012;18(1):89–92.

59. Keyser RJ, Lesage S, Brice A, Carr J, Bardien S. Assessing the prevalence of PINK1 genetic variants in South African patients diagnosed with early- and late-onset Parkinson’s disease. Biochem Biophys Res Commun. 2010;398(1):125–129.

60. Dekker MCJ, Suleiman JM, Bhwana D, et al. PRKN-related familial Parkinson’s disease: First molecular confirmation from East Africa. Parkinsonism Relat Disord. 2020;73:14–15.

61. Carr J, Guella I, Szu-Tu C, et al. Double homozygous mutations (R275W and M432V) in the ParkinGene associated with late-onset Parkinson’s disease. Mov Disord. 2016;31(3):423–425.

62. Bouhouche A, Tesson C, Regragui W, et al. Mutation Analysis of Consanguineous Moroccan Patients with Parkinson’s Disease Combining Microarray and Gene Panel. Front Neurol. 2017;8:567.

63. Gouider-Khouja N, Larnaout A, Amouri R, et al. Autosomal recessive parkinsonism linked to parkin gene in a Tunisian family. Clinical, genetic and pathological study. Parkinsonism Relat Disord. 2003;9(5):247–251.

64. Ibáñez P, Lesage S, Lohmann E, et al. Mutational analysis of the PINK1 gene in early-onset parkinsonism in Europe and North Africa. Brain. 2006;129(Pt 3):686–694.

65. Ishihara-Paul L, Hulihan MM, Kachergus J, et al. PINK1 mutations and parkinsonism. Neurology. 2008;71(12):896–902.

66. Lücking CB, Abbas N, Dürr A, et al. Homozygous deletions in parkin gene in European and North African families with autosomal recessive juvenile parkinsonism. The European Consortium on Genetic Susceptibility in Parkinson’s Disease and the French Parkinson’s Disease Genetics Study Group. Lancet. 1998;352(9137):1355-1356.

67. Norman BP, Lubbe SJ, Tan M, Warren N, Morris HR. Early Onset Parkinson’s Disease in a family of Moroccan origin caused by a p.A217D mutation in PINK1: a case report. BMC Neurol. 2017;17(1):153.

68. Zhu W, Huang X, Yoon E, et al. Heterozygous PRKN mutations are common but do not increase the risk of Parkinson’s disease. Brain. 2022;145(6):2077–2091.

69. Lesage S, Lohmann E, Tison F, et al. Rare heterozygous parkin variants in French early-onset Parkinson disease patients and controls. J Med Genet. 2008;45(1):43–46.

70. Yu E, Rudakou U, Krohn L, et al. Analysis of Heterozygous PRKN Variants and Copy-Number Variations in Parkinson’s Disease. Mov Disord. 2021;36(1):178–187.

71. Tishkoff SA, Reed FA, Friedlaender FR, et al. The genetic structure and history of Africans and African Americans. Science. 2009;324(5930):1035-1044.

72. Bryc K, Durand EY, Macpherson JM, Reich D, Mountain JL. The genetic ancestry of African Americans, Latinos, and European Americans across the United States. Am J Hum Genet. 2015;96(1):37–53.

73. Regier AA, Farjoun Y, Larson DE, et al. Functional equivalence of genome sequencing analysis pipelines enables harmonized variant calling across human genetics projects. Nat Commun. 2018;9(1):4038.

74. Iwaki H, Leonard HL, Makarious MB, et al. Accelerating Medicines Partnership: Parkinson’s Disease. Genetic Resource. Mov Disord. 2021;36(8):1795–1804.

75. Lange LM, Avenali M, Ellis M, et al. Elucidating causative gene variants in hereditary Parkinson’s disease in the Global Parkinson’s Genetics Program (GP2). NPJ Parkinsons Dis. 2023;9(1):100.

76. Harrison SM, Austin-Tse CA, Kim S, et al. Harmonizing variant classification for return of results in the All of Us Research Program. Hum Mutat. 2022;43(8):1114–1121.

77. Venner E, Muzny D, Smith JD, et al. Whole-genome sequencing as an investigational device for return of hereditary disease risk and pharmacogenomic results as part of the All of Us Research Program. Genome Med. 2022;14(1):34.

78. Van Hout CV, Tachmazidou I, Backman JD, et al. Exome sequencing and characterization of 49,960 individuals in the UK Biobank. Nature. 2020;586(7831):749-756.

79. Backman JD, Li AH, Marcketta A, et al. Exome sequencing and analysis of 454,787 UK Biobank participants. Nature. 2021;599(7886):628-634.

80. Bandres-Ciga S, Faghri F, Majounie E, et al. NeuroBooster array: A genome-wide genotyping platform to study neurological disorders across diverse populations. Mov Disord. Published online September 16, 2024. doi:10.1002/mds.29902

81. Vitale D, Koretsky M, Kuznetsov N, et al. GenoTools: An open-source Python package for efficient genotype data quality control and analysis. bioRxiv. Published online March 29, 2024. doi:10.1101/2024.03.26.586362

82. 1000 Genomes Project Consortium, Auton A, Brooks LD, et al. A global reference for human genetic variation. Nature. 2015;526(7571):68–74.

83. Siva N. 1000 genomes project. Nat Biotechnol. 2008;26(3):256.

84. Bray SM, Mulle JG, Dodd AF, Pulver AE, Wooding S, Warren ST. Signatures of founder effects, admixture, and selection in the Ashkenazi Jewish population. Proc Natl Acad Sci U S A. 2010;107(37):16222–16227.

85. Conley AB, Rishishwar L, Ahmad M, et al. Rye: genetic ancestry inference at biobank scale. Nucleic Acids Res. 2023;51(8):e44.

