## Supplementary Methods for "Large-scale genetic characterization of Parkinson’s disease in the African and African admixed populations" (from another version of this preprint: version 1, 2025-01-15)

Sequencing procedures

*GP2-BLAAC PD and NPDR Network*

Short-read whole genome sequencing (WGS) was performed by Psomagen (Rockville, MD, USA) using Illumina’s TruSeq DNA PCR-Free High Throughput Library Prep Kit (cat. # 20015963, Illumina, San Diego, CA, USA) ^1^. Sequencing was performed in 277 individuals from GP2-BLAAC PD and 168 individuals from the NPDR Network (108 PD cases and 60 controls).

The functional equivalence pipeline ^2^ from the Broad Institute was used to produce alignments and call small variants against a reference genome (GRCh38). We followed the quality metrics defined by the Accelerating Medicines Partnership Parkinson's Disease initiative (AMP-PD; <https://amp-pd.org>) ^3^ for sample-level WGS quality control. Joint-genotyped variants for all samples that passed quality control were generated from the Broad Institute's joint discovery pipeline. Only the high-quality variants flagged as “PASS” following variant quality score recalibration, with call rate >0.95, genotype quality >=20, read depth >=5, and heterozygous allele balance between 0.25 and 0.75 were retained, as described previously ^4^.

In addition to WGS, targeted sequencing was performed at Uniformed Services University of the Health Sciences (USUHS) in 383 additional GP2-BLAAC PD samples including 149 cases and 234 controls utilizing the Illumina TruSight One Sequencing Panel, which includes 4,800 genes and 12 Mb of genomic content (cat. 20025524 & 20029227, Illumina, San Diego, CA, USA). DNA samples were normalized to 50ng and quality control was performed with Qubit 4 Fluorometer. Seven samples did not pass quality control and were discontinued; 376 total samples were included for sequencing. Index, paired-end libraries at 150bp were produced using Illumina DNAPprep with Enrichment Tagmentation (cat # 20025524, Illumina, San Diego, CA, USA) and unique dual index sets (IDT for Illumina DNA/RNA UD Indexes Sets A, B, C, and D Tagmentation). Samples were pooled in batches of 96 samples and sequenced on the Illumina Novaseq6000. The sequencing data processing followed the same pipeline of WGS data as mentioned above. We performed joint-genotyping using Broad Institute's joint discovery pipeline and followed the same criteria for sample and variant quality control as for the WGS data.

*PD GENE*

Targeted-next generation sequencing data were utilized from the PD GENE dataset in GP2 release 8. Sequencing was performed by Fulgent Genetics, designed to examine coding regions and splicing junctions (https://www.fulgentgenetics.com/parkinson-disease-comprehensive). Details regarding the sequencing procedure for this dataset and data curation were described in Cook et al. 2024 ^5^.

*All of Us*

Participants were included from All of Us Controlled Tier Dataset release 7. Sequencing was performed by the Genome Centers funded by the All of Us Research Program ^6,7^. All genome centers used the same sequencing protocols that consisted of PCR-free 150 bp, paired-end libraries sequenced on the Illumina NovaSeq 6000 platform. Sequencing data was processed using DRAGEN v3.4.12 (Illumina, San Diego, CA, USA). The GRCh38 reference genome was used for alignment ^8^.

*UK Biobank*

Further details on whole-exome sequencing, variant calling, and quality control protocols can be found at Van Hout et al. 2020 and Backman et al. 2021 ^9,10^. In brief, samples were transferred to the Regeneron Genomics Center, where DNA libraries were created with mean sheared DNA fragment sizes of 200bp and unique asymmetric 10bp barcodes were added. Exome capture was performed using a variation of the IDT xGen Exome Research Panel v1.0, with two different lots being used between the first 50,000 samples and all other samples. DNA was amplified and quantified using qPCR before being sequenced using the Illumina NovaSeq 6000 platform with 75bp paired-end reads. S2 flow cells were used for the first 50,000 samples, while S4 flow cells were used for all remaining samples. Raw data were mapped to the GRCh38 reference genome using BWA-mem. Individual-level gVCF files were generated using the WeCall variant caller, which were joint-called into project-level pVCF files using GLnexus.

The rs3115534 intronic variant was obtained from UK Biobank’s whole-genome sequencing datasets. Whole-genome sequencing was conducted using the NovaSeq 6000 platform (<https://www.medrxiv.org/content/10.1101/2023.12.06.23299426v1>). These data were then analyzed with the DRAGEN v3.7.8 (Illumina, San Diego, CA, USA) software. Alignment was performed against the GRCh38 reference genome. Further details on quality control metrics can be found at https://biobank.ndph.ox.ac.uk/showcase/label.cgi?id=187.

Genotyping, quality-control and genetic ancestry estimation

The GP2-BLAAC PD samples were genotyped using the NeuroBooster (version 1.0; Illumina, San Diego, CA, USA) array platform ^11^. The array included 1,914,935 variants that encompass ancestry markers, identity by descent markers, and biological sex markers derived from X-chromosome single-nucleotide polymorphisms (SNPs). Out of these variants, 96,517 variants were customized based on neurodegenerative disease. The NPDR Network samples were genotyped on both the NeuroBooster and the NeuroChip arrays (version 1.0; Illumina).

Genotyping data generation, quality control, and ancestry predictions were conducted as follows: The machine learning method from the GenoTools pipeline (<https://github.com/GP2code/GenoTools>) was used to perform custom ancestry prediction and quality control on genotyping data according to previously described methods in Vitale et al. 2024 ^12^. Samples that failed quality control criteria were excluded based on call rate (<95%), sex determination inconsistency from the clinical data, or excess heterozygosity detection (|F| statistics > 0.15). Variants with Hardy-Weinberg Equilibrium P ≤ 1 × 10^-4^ in control samples were removed after the preliminary sample-level quality control was completed. Variants were further pruned to remove variants with non-random missingness first by using a cutoff of case-control status at P ≤ 1 × 10^-4^ and then by haplotype at P ≤ 1 × 10^-4^.

Ancestry estimates for the GP2-BLAAC PD, the NPDR Network, PD GENE, and UKB datasets were conducted by GenoTools (<https://github.com/GP2code/GenoTools>) using reference panels from the 1000 Genomes Project ^13^, Human Genome Diversity Project ^14^, and an Ashkenazi Jewish population ^15^. A standard protocol across all samples was used to perform ancestry estimates. In All of Us, principal components were generated from high-quality variants and projected onto HGDP and 1000 Genomes Project samples to estimate genetic ancestry using the Rye software as previously described in Conley et al. 2023 ^16^. While both African and African admixed reference panels were included in the ancestry estimation for the GP2-BLAAC PD, NPDR Network, PD GENE initiative and UKB, only the African reference panel was implemented for All of Us ancestry estimation. Ancestry inferences were applied to the 138 samples in the PD GENE dataset with available genotyping data while the remaining 145 samples were included based on their self-reported Black or African American race.

Copy number variant detection

Ninety-six cases from the GP2–BLAAC PD cohort were selected for CNV screening through a Multiplex Ligation-dependent Probe Amplification (MLPA) assay using the SALSA® MLPA® reagent kit (#EK1-FAM) and SALSA® MLPA® Probemix P051 (#P051-100R), according to the manufacturer’s instructions (MRC-Holland, Amsterdam, The Netherlands). Cases were prioritized for MLPA screening by family history of PD or an early age of onset (<50 years). Quantity of DNA was determined with a Qubit fluorometric dsDNA BR assay (#Q33326, Invitrogen, USA). DNA samples were diluted to 12.5 ng/uL in water for a total of 50 ng of DNA per sample and 1 µl of 50 mM Tris-HCl buffer at pH 8.5 (#BU-124S-85, Jena Biosciences, Germany) per reaction. PCR fragments were analyzed by capillary electrophoresis in an ABI 3730XL genetic analyzer (Applied Biosystems™, USA) using highly deionized (HiDi) formamide (#4311320, Applied Biosystems™, USA) and GeneScan™ 500 LIZ™ dye size standard (#4322682, Applied Biosystems™, USA). Data was analyzed using the Coffalyser.Net™ software package (MRC-Holland, Amsterdam, The Netherlands), according to the provided protocol.

1.	Rizig M, Bandres-Ciga S, Makarious MB, et al. Identification of genetic risk loci and causal insights associated with Parkinson’s disease in African and African admixed populations: a genome-wide association study. *Lancet Neurol*. 2023;22(11):1015-1025.

2.	Regier AA, Farjoun Y, Larson DE, et al. Functional equivalence of genome sequencing analysis pipelines enables harmonized variant calling across human genetics projects. *Nat Commun*. 2018;9(1):4038.

3.	Iwaki H, Leonard HL, Makarious MB, et al. Accelerating Medicines Partnership: Parkinson’s Disease. Genetic Resource. *Mov Disord*. 2021;36(8):1795-1804.

4.	Lange LM, Avenali M, Ellis M, et al. Elucidating causative gene variants in hereditary Parkinson’s disease in the Global Parkinson's Genetics Program (GP2). *NPJ Parkinsons Dis*. 2023;9(1):100.

5.	Cook L, Verbrugge J, Schwantes-An TH, et al. Parkinson’s disease variant detection and disclosure: PD GENEration, a North American study. *Brain*. 2024;147(8):2668-2679.

6.	All of Us Research Program Investigators, Denny JC, Rutter JL, et al. The “All of Us” Research Program. *N Engl J Med*. 2019;381(7):668-676.

7.	Harrison SM, Austin-Tse CA, Kim S, et al. Harmonizing variant classification for return of results in the All of Us Research Program. *Hum Mutat*. 2022;43(8):1114-1121.

8.	Venner E, Muzny D, Smith JD, et al. Whole-genome sequencing as an investigational device for return of hereditary disease risk and pharmacogenomic results as part of the All of Us Research Program. *Genome Med*. 2022;14(1):34.

9.	Van Hout CV, Tachmazidou I, Backman JD, et al. Exome sequencing and characterization of 49,960 individuals in the UK Biobank. *Nature*. 2020;586(7831):749-756.

10.	Backman JD, Li AH, Marcketta A, et al. Exome sequencing and analysis of 454,787 UK Biobank participants. *Nature*. 2021;599(7886):628-634.

11.	Bandres-Ciga S, Faghri F, Majounie E, et al. NeuroBooster array: A genome-wide genotyping platform to study neurological disorders across diverse populations. *Mov Disord*. Published online September 16, 2024. doi:[10.1002/mds.29902](http://dx.doi.org/10.1002/mds.29902)

12.	Vitale D, Koretsky M, Kuznetsov N, et al. GenoTools: An open-source Python package for efficient genotype data quality control and analysis. *bioRxiv*. Published online March 29, 2024. doi:[10.1101/2024.03.26.586362](http://dx.doi.org/10.1101/2024.03.26.586362)

13.	1000 Genomes Project Consortium, Auton A, Brooks LD, et al. A global reference for human genetic variation. *Nature*. 2015;526(7571):68-74.

14.	Siva N. 1000 genomes project. *Nat Biotechnol*. 2008;26(3):256.

15.	Bray SM, Mulle JG, Dodd AF, Pulver AE, Wooding S, Warren ST. Signatures of founder effects, admixture, and selection in the Ashkenazi Jewish population. *Proc Natl Acad Sci U S A*. 2010;107(37):16222-16227.

16.	Conley AB, Rishishwar L, Ahmad M, et al. Rye: genetic ancestry inference at biobank scale. *Nucleic Acids Res*. 2023;51(8):e44.
