## Supplementary material for "Large-scale genetic characterization of Parkinson’s disease in the African and African admixed populations": Figure legends (from another version of this preprint: version 1, 2025-01-15)

**Figure 1. Demographic and clinical characteristics of datasets under study.**

The figure illustrates distributions of age, sex, and the number of cases and controls across five datasets in this study: All of Us, Black African and African American Connections to Parkinson's Disease Study (BLAAC PD) as a part of the Global Parkinson’s Genetics Program (GP2), Nigerian Parkinson’s Disease Research (NPDR) Network, the PD GENEration (PD GENE) initiative and UK Biobank (UKB). Both African and African admixed reference panels were included in the ancestry estimation for the BLAAC PD, Nigerian Parkinson’s Disease Research Network, PD GENE initiative and UKB, while only African reference panel was implemented for All of Us ancestry estimation.

**Figure 2.** **Workflow**

Our workflow begins with creating cohorts within the datasets. We utilized short-read sequencing data to characterize Parkinson’s disease, parkinsonism and related neurodegenerative disease genes. We prioritized missense, frameshift, start loss, stop loss, stop gain, and splicing variants with a minor allele frequency < 0.01 and CADD score > 12.37.

**Figure 3.** **Distribution of variants in Parkinson’s disease and parkinsonism-related genes.**

Parkinson’s disease and parkinsonism related genes included for analysis. †het indicates autosomal recessive inheritance. “Very high” indicates genes with very high confidence being related to disease, “High” indicates high confidence, and “Low” indicates low confidence based on replication in the literature (Blauwendraat et al., 2020). Five cohorts were included: Black African and African American Connections to Parkinson's Disease Study (BLAAC PD), as a part of the Global Parkinson’s Genetics Program (GP2), Nigerian Parkinson’s Disease Research (NPDR) Network, the PDGENEration initiative, the All of Us Program and the UK Biobank (UKB). The width of the connecting line between the cohort and each gene is correlated to the number of variants found.

**Figure 4. Prioritized rare protein-altering or splicing variants in known autosomal dominant, recessive and genetic risk factors for Parkinson’s disease**

Autosomal recessive genes for PD include: *PRKN, PINK1, DJ-1*, and *VPS13C*. Genetic risk factors for PD include: *GBA1* and *LRRK2*. Autosomal dominant genes for PD include: *SNCA* and *VPS35*. For the latter group, these genes were not individually plotted since only one variant in each autosomal dominant gene was detected. Rare variants were included if minor allele frequency (MAF) was lower than 0.01 and CADD score > 12.37. Novel indicates variants not previously reported in the literature. Likely benign and benign variants, as well as likely pathogenic and pathogenic variants according to pathogenicity predictors, are presented together for each gene. VUS=variant of uncertain significance. (Novel variants in other PD genes include two in *DNAJC6*, one in *SNCA*, two in *VPS35*, and one in *SYNJ1*).

**Figure 5. Distribution of age at onset among Parkinson’s disease prioritized variant carriers**

**Figure 6. Mutation sites of identified genetic variants on the proteins encoded by Parkinson’s disease-associated genes**

Identified genetic variants were mapped on AlphaFold-predicted protein structures encoded by eight genes associated with Parkinson’s disease (*DJ-1, GBA1, LRRK2, PINK1, PRKN, SNCA, VPS13C, VPS35*).

**Figure 7. Biallelic PRKN deletion identified in an early-onset Parkinson’s disease case.**

A) Multiplex Ligation-dependent Probe Amplification (MLPA) was performed in early-onset Parkinson’s disease cases and/or individuals with positive family history. One early-onset Parkinson’s disease with an age at onset of 27 years was detected to have a decrease in the height of the electropherogram peak at 202 and 477 nucleotides (red arrows), which correspond to *PRKN*-4 and *PRKN*-3 probes, respectively. B) The probe ratios indicate a *PRKN* exon 3 heterozygous deletion (0.5 ratio) and a *PRKN* exon 4 homozygous deletion (0 ratio).

**Supplementary Figure 1. Ancestry prediction clustering for each cohort under study**

Three dimensional principal components analysis to group individuals based on their genetic makeup. A) UK Biobank (UKB) B) All of Us C) GP2-BLAAC PD D) Nigerian Parkinson’s Disease Research Network E) PD GENEration. Blue color indicates African ancestry, and green color indicates African admixed ancestry.

**Supplementary Figure 2. Candidate variants identified in Parkinson’s disease genes in individuals of African and African admixed ancestry.**

*red highlighted: variants previously reported as pathogenic or likely pathogenic. For the recessive PD genes, variants only found in early-onset cases were included.

**Supplementary Figure 3. *PRKN* B allele frequencies and log-R ratios from genotyping data.**

Copy number variants (CNV) in *PRKN* were screened in 1,167 PD cases. A total of 872 African and 295 African admixed ancestry individuals were included in this study. Eight samples had positive CNV results for *PRKN* as shown in A) indicating a heterozygous deletion in exon 3, B) a heterozygous duplication in exon 2, C) a heterozygous deletion in exons 3 and 4 and a second potential heterozygous deletion in exon 4, D) a homozygous deletion in exons 3 and 4, E) a homozygous deletion in exons 3 and 4, F) a heterozygous duplication in exon 1, heterozygous deletion in exon 10, and a region of homozygosity in exons 2 and 3; G) a heterozygous duplication in exon 2, and H) a heterozygous deletion in exons 3 and 4. Reference Table 4 for demographic information.
